# Transcriptome-wide association study of Alzheimer disease reveals many differentially expressed genes and multiple biological pathways in brain tissue from African American donors

**DOI:** 10.1101/2024.10.29.24316311

**Authors:** Mark W. Logue, Adam Labadorf, Nicholas K. O’Neill, Dennis W. Dickson, Brittany N. Dugger, Margaret E. Flanagan, Matthew P. Frosch, Marla Gearing, Lee-Way Jin, Julia Kofler, Richard Mayeux, Ann McKee, Carol A. Miller, Melissa E. Murray, Peter T. Nelson, Richard J. Perrin, Julie A. Schneider, Thor D. Stein, Andrew F. Teich, Juan C. Troncoso, Shih-Hsiu Wang, Benjamin Wolozin, Jesse Mez, Lindsay A. Farrer

## Abstract

**Background:** The genetic basis of Alzheimer disease (AD) in African American (AA) individuals is much less well understood than in European-ancestry (EA) individuals. Furthermore, relatively few AA donors have been included in postmortem AD studies.

**Methods:** We generated transcriptome-wide bulk-level gene expression data derived from pre-frontal cortex (PFC) tissue obtained from 179 AA brain donors with a pathological diagnosis of AD or control. This sample was augmented by previously generated RNAseq data derived from PFC tissue from another group of 28 AA donors, yielding a total sample of 125 AD cases and 82 neuropathologically determined controls who were enrolled at 12 AD research centers across the United States. Expression of 33,611 genes was compared between AD cases and controls using LIMMA including covariates for age, sex, cell-type frequencies, and RNA integrity number. A false discovery rate corrected p-value (padj) was used to account for multiple testing. Gene-ontology (GO) term enrichment analysis was performed using GOseq.

**Results:** Transcriptome-wide significant differential expression was observed with 482 genes among which the most significant, *ADAMTS2,* showed 1.52 times higher expression in AD cases compared to controls (p=2.96×10^−8^, padj=0.0010). Many of the differentially expressed genes are involved in mitochondrial energy production. Examination of differentially expressed genes observed previously in brain tissue from EA AD cases and controls revealed that 385 (35%) were nominally significant, 65 (5.8%) were significant after multiple test correction, and most (89%) had the same effect direction in the AA sample. Several other significant associations were not observed in the EA cohort, and these genes may be especially relevant to AD risk in the AA population (e.g., *EFR3B*, *IRS4*, and *CA12*). Examination of potential regulatory effects of AA GWAS-implicated AD risk variants identified several expression quantitative trait loci.

**Conclusions:** In this first large-scale transcriptome-wide gene expression study of AD in brain from AA donors, we identified many differentially expressed genes, including *ADAMTS2* which was recently reported to be differentially expressed in brain between pathologically confirmed EA AD cases with and without AD clinical symptoms. These results represent a substantial advance in knowledge about the genetic basis of AD in the AA population and suggest promising new targets for treatment.

## Introduction

Alzheimer disease (AD) is the most common form of dementia and a major public health concern, with 1 in 45 Americans affected by 2050 unless effective treatments and/or interventions are developed and implemented^1,2^. The prevalence of AD is about two times higher in the Black/African American (AA) population compared to White/European-ancestry (EA) individuals living in the United States^3^. Some of this difference is due to social determinants of health such as disparities in health care access and quality of education^4–6^, biases in testing^7,8^, and higher rates of AD risk factors such as cardiovascular disease^9^ and diabetes^10^ in those who identify as AA, although the causes of these differences are not completely known^11^. Group differences have also been noted in the pattern of association with AD risk variants^12,13^. For example, *APOE* ε4 is more frequent in the AA population, but the reported AD risk associated with *APOE* ε4 heterozygosity and especially homozygosity is smaller in AA compared to EA cohorts^14^.

Much of our understanding of the genetic basis of AD in AA individuals is derived from candidate gene and genome-wide association studies, although with much smaller sample sizes than those included in genetic studies of EA cohorts^15^. For example, a recent large AD GWAS in EA cohorts including approximately 85,000 AD cases and 296,000 controls as well as information about proxy AD (reported parental dementia) cases identified genome-wide significant associations with 75+ loci^16^. By comparison, the largest GWAS of AD and AD-related dementia (ADRD) in AA cohorts^17^ included 4,012 ADRD cases, 18,435 controls and 52,611 AD proxy cases and controls from the Million Veteran Program (MVP)^18^ and an additional 2,784 AD cases and 5,222 controls assembled by the Alzheimer’s Disease Genetics Consortium (ADGC)^19^. That study confirmed associations previously established in EA studies (i.e. *APOE*, *ABCA7*, *CD2AP*, and *TREM2*) and identified highly significant novel associations with variants in *ROBO1* and *RP11-340A13.2*. In addition, associations have been identified with rare pathogenic variants in AA cohorts that are rare or absent in EA populations, suggesting that some AD-related mechanisms may be more easily discernable in particular ancestry groups^12,23,24^.

Although many studies have examined differential gene expression (DGE) in brain tissue from AD cases and controls in EA or mixed ancestry cohorts using bulk tissue, single-cell sequencing, and individual cell types (e.g. ^20–24^), the number of AA individuals in these studies was unspecified or too small to identify significant findings within this group alone. One study identified AA-specific gene expression associations from analysis of RNA sequencing (RNA-seq) data obtained from peripheral blood samples from 231 AA participants and 234 EA participants^25^. In this study, we evaluated RNA-seq data derived from post-mortem prefrontal cortex (PFC) tissue from 207 AA brain donors (125 pathologically confirmed AD cases and 82 controls). The most significantly differentially expressed gene, *ADAMTS2*, was also one of the top-ranked genes in a concurrent study comparing gene expression in brain tissue from EA AD cases and controls^26^, and the most significant differentially expressed gene between the pathologically confirmed AD cases with and without clinical AD symptoms prior to death^27^. We also found evidence suggesting modulation of gene expression as a potential mechanism for some AD risk variants implicated in a recent AA AD/dementia GWAS^17^.

## Methods

### Participants, specimens and diagnostic procedures

We obtained frozen brain tissue specimens from 229 donors identified as AA individuals from 13 Alzheimer’s Disease Research Center (ADRC) brain banks: (1) Boston University ADRC (BU), (2) Goizueta ADRC at Emory University (GADRC), (3) Johns Hopkins ADRC (JH), (4) University of Kentucky Alzheimer’s Disease Center Tissue Bank (UKY), (5) Florida Brain Bank and ADRC at the Mayo Clinic (Jacksonville, FL), (6) Rush Alzheimer’s Disease Core Center (RUSH), (7) University of Pittsburgh ADRC (PITT), (8) Carroll A. Campbell, Jr. Neuropathology Laboratory at the Medical University of South Carolina (USC), (9) The Charles F. and Joanne Knight ADRC at Washington University in Saint Louis (WASHU), (10) Massachusetts ADRC (MGH), (11) University of California-Davis ADRC (UCD), (12) Bryan Brain Bank and Biorepository at the Duke-UNC ADRC (DUKE), and (13) the Northwestern ADC Neuropathology Core (NW). In addition, we received previously generated RNAseq data for 28 donors (18 cases and 10 controls) from the Columbia University ADRC (CU). Scores measuring the severity of neurofibrillary tangle involvement (Braak) and neuritic amyloid deposition (CERAD) scores obtained from the brain banks were harmonized using the ADRC data dictionary to the standard 0-6 coding for Braak and 0-3 coding for CERAD. We used National Institute on Aging – Alzheimer Association guidelines^28^ and standard NIA Reagan scoring criteria to classify the donors as AD cases (intermediate likelihood or high likelihood of AD) and controls (not AD or low likelihood of AD).

### RNA sequencing data generation and quality control

RNA was extracted from the PFC tissue using the Maxwell® RSC simplyRNA Tissue Kit from Promega according to the manufacturer’s instructions. RNA integrity number (RIN) values were assessed using Agilent 2100 Bioanalyzer RNA Chip (Santa Clara). RNAseq was performed on samples with RIN scores > 2 by the Yale Center for Genome Analysis in two batches of 90 and 99 samples, respectively. RNAseq data were successfully generated for all but one sample. Initial QC was performed using FastQC (http://www.bioinformatics.babraham.ac.uk/projects/fastqc/). Adapter trimming and removal of low-quality reads was performed using Trimmomatic^29^ based on the Trueseq 2 Universal Adapter reference sequence. BAM files containing paired-end RNAseq read data were obtained for the CU participants. Generation of RNAseq data for the CU donors is described in detail elsewhere^30^. Reads were extracted using BEDTools^31^ and aligned to the genome jointly with the other cohorts as follows. Paired-end reads were aligned to the hg38 human reference genome using STAR^32^. Quantification at the gene and transcript level was performed using RSEM^33^. Post-alignment QC was performed using RSeQC^34^, and aggregation of the QC across samples was performed using MultiQC^35^. Two samples with mapping rates < 50% were excluded from subsequent analysis. All of the remaining samples had mapping rates > 80%. A principal component analysis of the rLog samples using R did not reveal any outliers (<6 SD), but examination of the PCs indicated clustering by batch (Batch 1, Batch 2, CU), and hence batch was included as a covariate in the differential expression analyses. Sex concordance was checked in by examining the total number of reads mapped to 4 Y-chromosome genes: *KDM5D*, *DDX3Y*, *USP9Y*, and *UTY*. We identified one mismatch, which was excluded. Cell-type estimates for use as covariates were generated using the DeTREM method^36^ with single-nuclei reference data from a prefrontal cortex AD study^37^.

### Genotype data

DNA was extracted from brain tissue using Maxwell RSC Blood DNA capture kits l. Samples were sent to The Children’s Hospital of Philadelphia Center for Applied Genomics for genotyping using the Illumina Global Diversity Array (∼1.8 million variants). Post-genotyping QC was performed using plink v1.90b3.36 and v2.00a2.3^38^. Two samples were excluded for high missing rates (>5%). X-chromosome heterozygosity was examined to identify potential sex mismatches. Two samples were removed due to discordant sex. Imputation of variants not on the array was performed using the Michigan Imputation Server (https://imputationserver.sph.umich.edu/) based on 1000 Genomes phase 3v5 reference data^39^. After cleaning, genotype data, valid RNAseq data, and non-missing covariates were available for 177 donors.

### Differential Gene Expression Analysis

DGE analysis was performed for genes with ≥1 read in half of the sequenced cohort using DEseq2^40^. A Benjamini Hochberg False discovery rate corrected p-value (padj, often called a q-value)^41,42^ was computed to adjust for 33,611 genes examined. Regression models included covariates for age at death, sex, batch, RIN, and the estimated proportions of inhibitory neurons, excitatory neurons, microglia, oligodendrocytes, astrocytes, and endothelial cells. Post-mortem interval (PMI) was not included as a covariate because this information was missing for 20 samples, however, we did perform sensitivity analyses to confirm that excluding PMI did not substantially bias the results. Volcano plots were generated using the EnhancedVolcano R package (https://github.com/kevinblighe/EnhancedVolcano). Expression differences are reported as log2 fold change (L2FC).

### Transethnic Comparisons of DGE and Genetic Association Findings

First, we examined differential expression of *APOE and* genes implicated in EA AD GWAS^16^, AA dementia GWAS^17^, and genes implicated in other AA AD genetic studies^15^ at an FDR corrected significance level that adjusted for the number of distinct genes examined within each gene set. We also assessed by linear regression potential regulatory effects of variants having a minor allele frequency ≥5% and associated with dementia risk in AA cohorts (Table 2) on expression of genes within 200 kb of the variant according to the ensemble v 75 (GRCH37) database accessed via the R biomaRt library. The model included covariates for age at death, sex, batch, RIN, and the estimated cell-type proportions. FDR-corrected p values were calculated for each variant adjusting for the number of genes examined. Top-ranked DGE results from the AA cohort were compared to those obtained from frontal cortex tissue of 526 autopsy-confirmed AD cases and 456 controls from four EA cohorts^43^ using a Χ^2^ test.

### Overrepresentation and Gene Network Analyses

Overrepresentation analysis was conducted to identify GO terms (biological processes and pathways) that are enriched for the top-ranked DEGs using GOseq^44^, a method that adjusts for any differential likelihood of significance due to gene size. To increase interpretability and limit multiple testing, we only tested for overrepresentation within GO Biological Process and GO Molecular Function categories. Prior to performing gene network analyses, RNA-seq data were corrected using COMBAT-SEQ^45^ to adjust for batch effects but preserve age, sex, and AD effects across the batches. The batch-corrected counts were normalized and log transformed using the DEseq2^40^ package, and networks of correlated genes were identified by analysis of the resulting rLog values using Weighted Gene Co-Expression Network Analysis (WGCNA)^46^. This analysis was performed using the blockwiseModules function in the WGNA R package to generate signed networks with power=15 and options cut height=0.75, min module of 30, max block of 4000, reassign threshold=0, mergeCutHieght=0.25, and the rest of the options left at the default. Only highly expressed genes (>10 reads in half of the cohort) were included in this analysis to reduce computational complexity.

## Results

After excluding 34 samples that were obtained from non-PFC regions or had RNA quality or sequencing issues, and 1 sex mismatched sample, 212 participants (including 82 controls, 125 AD cases and 5 subjects lacking neuropathological data to determine AD status) remain for subsequent analyses (**Table 1**). Participants with unknown neuropathological AD status were excluded from the DGE analysis but included in eQTL analysis. The sex ratio is approximately 1 among the controls, but a higher proportion of cases are women. The mean age at death was 5.3 years greater in AD cases than controls (p=0.0040).

**Table 1:**
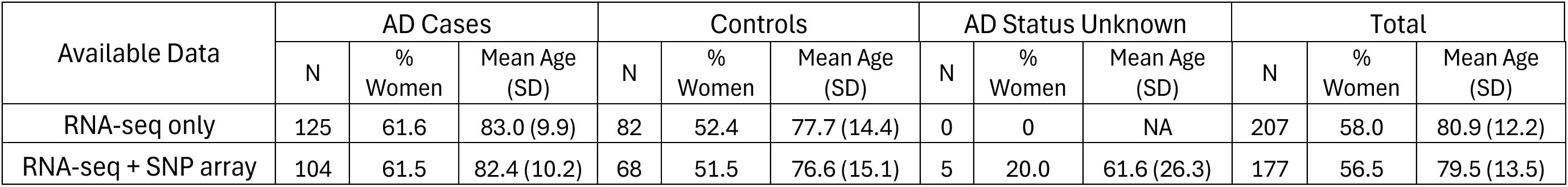
Subject characteristics.

### Genes Differentially Expressed Between AD Cases and Controls

Transcriptome-wide significant (TWS) differences were observed for 482 of the 33,611 sufficiently expressed genes, among which 174 had higher and 308 had lower expression in AD cases than controls (**Figure 1, Supplementary Table 1**). The most significant DEG, *ADAMTS2*, was expressed 1.52 times higher in AD cases than controls (L2FC=0.60, p=2.96×10^−8^, **Table 2, Supplementary Figure 1**). Sensitivity analyses showed that excluding PMI as a covariate and excluding younger (age <60 at time of death) donors did not substantively impact the results (**Supplementary Figure 2, Supplementary Table 2**). By comparison, none of the AD loci established by the largest GWAS in EA and AA cohorts^16,17^ were significant DEGs after multiple test correction, but expression of *IG1FR*, which was implicated in an AA AD GWAS (Kunkle 2021)^19^ due to a rare variant association (rs570487962), was significantly higher in AD cases (L2FC=0.11, p=0.0013, padj=0.019, **Supplementary Table 3**).

**Figure 1:**
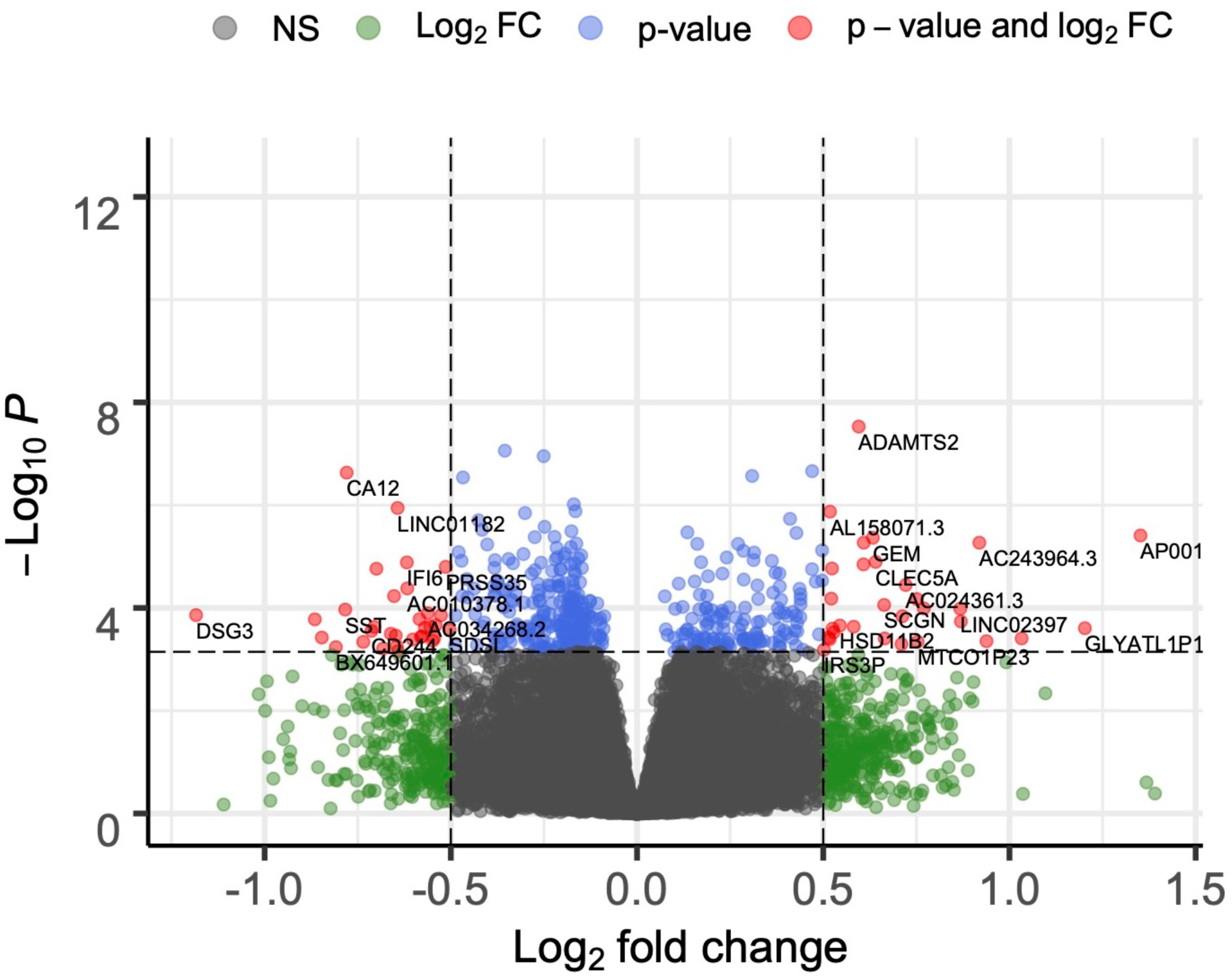
Volcano plot of a transcriptome-wide differential gene expression analysis of 125 neuropathologically determined African American AD cases and 82 neuropathologically determined African American controls. The color indicates whether or not genes passed multiple testing corrected significance thresholds (p-value) and whether or not L2FC >0.05, or both.

**Table 2:** 20 genes most significantly differentially expressed between African American AD cases and controls.

| Gene | L2FC | P | p<sub>adj</sub> |
| --- | --- | --- | --- |
| *ADAMTS2* | 0.60 | 2.96E-08 | 0.0010 |
| *AP002360.1* | -0.35 | 8.71E-08 | 0.0012 |
| *EFR3B* | -0.25 | 1.11E-07 | 0.0012 |
| *IRS4* | 0.47 | 2.18E-07 | 0.0014 |
| *CA12* | -0.78 | 2.33E-07 | 0.0014 |
| *ITPKB* | 0.31 | 2.70E-07 | 0.0014 |
| *PDE10A* | -0.47 | 2.89E-07 | 0.0014 |
| *TDRKH* | -0.17 | 9.62E-07 | 0.0040 |
| *LINC01182* | -0.64 | 1.13E-06 | 0.0040 |
| *TFCP2* | -0.17 | 1.31E-06 | 0.0040 |
| *AL158071.3* | 0.52 | 1.34E-06 | 0.0040 |
| *EHHADH* | -0.30 | 1.42E-06 | 0.0040 |
| *PIEZO2* | 0.41 | 1.84E-06 | 0.0047 |
| *P2RY1* | -0.43 | 1.97E-06 | 0.0047 |
| *AC018647.2* | -0.25 | 2.64E-06 | 0.0059 |
| *NOX4* | -0.42 | 3.02E-06 | 0.0062 |
| *ECHS1* | -0.18 | 3.21E-06 | 0.0062 |
| *ZCCHC14* | 0.14 | 3.38E-06 | 0.0062 |
| *LINC01094* | 0.43 | 3.48E-06 | 0.0062 |
| *AP001180.1* | 1.35 | 3.89E-06 | 0.0063 |

L2FC = log<sub>2</sub> fold change

Trans-ethnic comparison revealed that four of the most significant DEGs in the AA group (*ADAMTS2*, *ITPKB*, *TDRKH*, and *LINC0194*) were also differentially expressed between AD cases and controls at a TWS level in a large EA sample^26^.

Additionally, the most significant DEG in the EA study, *EMP3*, was also significant in the AA cohort with a similar effect direction (L2FC=0.33, p=0.00037). The overlap in TWS associations was more than would be expected by chance (p< 2.2×10^−16^), with 6% of TWS DEGs in the EA sample also TWS in the AA cohort (65/1092) vs 1.4% TWS significant in the overall AA cohort. Importantly, the direction of effect is the same for the 65 DEGs that are TWS in the AA and EA cohorts.

### Enrichment of DEGs involved in mitochondrial function, phosphorylation, DNA binding, and transcription

Enrichment analysis of the differentially expressed genes yielded 71 significant Enriched GO terms, especially those involving oxidative reduction processes and mitochondrial energy generation. All of the top 10 GO terms include 11 nuclear-encoded NDUF (NADH:ubiquinone oxidoreductase core subunit) genes (*NDUFA1*, *NDUFA2*, *NDUFA3*, *NDUFA6*, *NDUFA7*, *NDUFB1*, *NDUFB2*, *NDUFB3*, *NDUFB8*, *NDUFS3*, and *NDUFS5*) and cytochrome c oxidase genes (*COX15*, *COX4I1*, *COX6B1*) which were all expressed less in AD cases compared controls **(Supplementary Table 4**). Because *NADH-* and *COX*-gene encoded molecules form parts of the mitochondrial respiratory chain, we also examined but did not find differential expression of 16 mitochondrial DNA genes (p>0.07). Gene network analysis identified 27 networks ranging in size from 44 to 2,944 genes. Eigengenes (PCs representing gene activity) for three networks (lightyellow, magenta, salmon) remained significant after correction for multiple testing (**Supplementary Table 4**). The lightyellow network included 111 genes and was enriched for those involved in regulation of developmental processes or phosphorylation. Expression for most (7/9) of the AD-associated (p<0.05) genes in this network was higher in AD cases than controls. The magenta network was enriched for genes involved in DNA binding, transcription regulation, and cilium organization. Salmon was the most significantly associated network with AD (p=5.19×10^−6^, pcor=0.00014) and was enriched for genes related to protein complexes, oxidative phosphorylation, and metabolic processes.

### eQTLs among AD-associated variants identified by GWAS in AA cohorts

SNP association and gene expression data were available for 177 AA brain donors. Ten of 12 SNPs with a MAF > 0.05 which were significantly associated with AD/ADRD in the large AA GWAS^17^ were located within 200 kb of at least one gene (range 1 to 15, mean=5.5). After correction for multiple testing, significant eQTL associations were observed with five variants (**Table 3**). The minor alleles of two highly correlated SNPs (rs2234253 and rs73427293) in the *TREM2*/*TREML2* region (D’=1, R^2^=0.99 in the 1000 Genomes AFR reference data) were associated with higher expression of *ADCY10P1* (p=0.026) and *TREML2* (p=0.044). The minor allele of *CD2AP* SNP rs7738720 was associated with reduced *CD2AP* expression (p=0.026). The minor allele of *MSRA* intronic SNP rs4607615 was associated with expression of *MSRA* (p=0.026), *RP1L1* (p=0.041), and most strongly with *PRSS51* (p=6.11×10^−7^) even though rs4607615 is 61 kb downstream of that gene. The minor allele *APOE* SNP rs429358 which encodes the ɛ4 isoform is associated with reduced expression of *APOE* (p=5.61×10^−4^) and adjacent genes *APOC2* and *CLPTM1* (p=0.017 for both).

**Table 3:** Associations of AA GWAS-implicated variants and expression of nearby (<200 kb) genes.

| SNP | gene | Direction* | P | P<sub>cor</sub> |
| --- | --- | --- | --- | --- |
| **rs2234253** | *ADCY10P1* | + | 0.0048 | 0.0263 |
|  | *TREML2* | + | 0.0038 | 0.0263 |
| **rs73427293** | *ADCY10P1* | + | 0.0080 | 0.0437 |
|  | *TREML2* | + | 0.0044 | 0.0437 |
| **rs7738720** | *CD2AP* | - | 0.013 | 0.026 |
| **rs4607615** | *MSRA* * | - | 0.026 | 0.041 |
|  | *RP1L1* | + | 0.031 | 0.041 |
|  | *PRSS51* | - | 1.53E-07 | 6.11E-07 |
| **rs429358** | *APOE* * | - | 5.61E-04 | 8.42E-03 |
|  | *APOC2* | - | 0.0033 | 0.0166 |
|  | *CLPTM1* | - | 0.0023 | 0.0166 |

\*Direction of the effect of each minor allele on gene expression.

## Discussion

We performed the largest to date transcriptome-wide study of AD using brain tissue from AA donors. This is an important step in deciphering the genetic architecture and underlying mechanisms of AD risk in this population, in light of evidence that nearly all of the established AD risk variants are population-specific or have divergent frequencies across populations^17,19,47,48^. Additionally, because exposure to some AD risk factors differ between White and Black Americans including those that disproportionally affect the Black population in the US (e.g., vascular disease, diabetes, low education, and others linked to diet, income and occupation)^15^, the degree to which some biological pathways lead to AD likely varies across populations. In spite of such expected differences, we found high correspondence of differential expression of many genes in the pre-frontal cortex from AA and EA brain donors.

*ADAMTS2* was the most significant DEG in our study and among the top-ranked DEGs assessed in prefrontal cortex tissue in a very recent study of 982 EA brain donors (p=5.1×10^−13^)^49^ with the same effect direction in both populations. In addition, the Li et al. study^27^ found that *ADAMTS2* was the most significant DEG between AD cases who had an antemortem clinical AD diagnosis versus AD cases who were cognitively healthy prior to death (i.e., cognitively resilient). Taken together, these findings suggest that lower *ADAMTS2* expression reduces the risk of cognitive impairment among those who develop hallmark AD pathology. *ADAMTS2* is a member of the disintegrin and metalloproteinase with thrombospondin motifs (*ADAMTS*) family of genes that perform a diverse set of functions^49^. The most well studied role of ADAMTS2 relates to its proteolytically processed form resulting in a procollagen N-proteinase. *ADAMTS2* knockout mice have progressive skin fragility and male sterility^50^. In humans, *ADAMTS2* mutations cause a recessive form of Ehlers-Danlos syndrome^51^. Other studies have implicated ADAMTS2 in neurological processes related to AD risk. Yamakage et al. demonstrated in *ADAMTS2* knockout mice that ADAMTS2 induces cleavage of Reelin in the prefrontal cortex and hippocampus, impairing its function^52^. Mouse studies suggest Reelin is protective and found that its level is negatively associated with tau phosphorylation and amyloid plaque formation^53,54^. A recent study of resilience in a 67 year-old cognitively intact individual carrying a highly penetrant *PSEN1* mutation that causes autosomal dominant early onset AD identified a putatively protective gain of function variant (H3447R) in *RELN* (the gene encoding reelin), which contributed to reduced Tau phosphorylation in mouse AD models^55^. Results of human studies of Reelin levels and *RELN* expression in brain tissue are less consistent, showing increased^53,56^, reduced^54,57^, or no difference^58^ in reelin levels and RELN expression in AD vs control tissue. In this cohort, we did not observe an association of *RELN* expression with AD in PFC tissue (p=0.35). Nevertheless, our observation of higher expression of *ADAMTS2* in PFC from AD cases and its role in the Reelin pathway supports the hypothesis that *ADAMTS2* inhibition might be an AD therapeutic strategy^52^.

Four of the other six most significant (p<3.0×10^−7^) DEGs (*ITPKB, IRS4, CA12,* and *PDE10A*) were previously implicated in AD-related processes. *ITPKB* expression was shown to be higher in AD cases than controls^59^. Mouse studies have linked ITPKB activation and higher *ITPKB* expression to AD pathology.^60,61^ *IRS4* which encodes insulin receptor substrate 4 is known to function in several processes relevant to AD including interacting with endosomes to control Aβ levels in neurons^62^. In addition, insulin signaling and resistance, and diabetes more generally, have been strongly implicated in AD risk (e.g. ^63–66^). *Irs4* expression was reduced in B6.*APB^Tg^* mice in the early stages of AD pathology^62^, which does not match our observation of higher *IRS4* expression in AD cases, but this may be due to the later stage of disease in the post-mortem human brain. IRS-4 interacts with IRS-2^67^, which is also among the TWS DEGs in our AA study, and disruption of IRS-2 impacts tau phosphorylation, although the direction of effect is not consistent across studies^68,69^.

*CA12* is part of the carbonic anhydrase (CA) family which has been linked to learning and memory^70^*. CA12* expression in the caudate nucleus was identified as regulated by rs117618017, a SNP in the neighboring gene *APH1B*^71^ which was robustly associated with AD risk in GWAS^16,72^. Repurposing existing FDA-approved CA inhibitors has been suggested as a potential dementia treatment^73^. PDE10A is member of the phosphodiesterase family of enzymes involved in the breakdown of cAMP and/or cGMP^74^. PDE inhibitors have been shown to increase memory task performance in animals (e.g. ^75,76^) and have been investigated as potential treatments for autism spectrum disorder^77^, schizophrenia^78^, and Parkinson disease^79,80^. A clinical trial of a PDE1 inhibitor in AD cases did not find a benefit^81^, but other PDE inhibitors may have efficacy^74^. Here, we found that *PDE10A* expression is lower in AD brain tissue, which is perhaps counter to the notion that PDE10A inhibition is an effective treatment strategy. However, prior expression studies of multiple PDE genes yielded conflicting results^74^.

Several other TWS DEGs have strong connections to AD. *PSENEN* encodes a subunit of the gamma-secretase complex including presenilin^82^. *ROBO3* is a homolog of *ROBO1* which emerged as a genome-wide significant locus in an AA AD GWAS^16^. *HDAC4* is one of several histone deacetylases previously implicated in AD and identified as potential targets for AD treatment^83^.

Analyses of pathways enriched for significant DEGs highlighted the importance of many nuclear genes involved in mitochondrial energy production, including 3 cytochrome c oxidase genes and 11 genes encoding *NUDF* subunits, all of which were expressed at a lower level in AD cases than controls. NDUF subunits form part of mitochondrial respiratory complex I, a multimeric enzyme composed of 44 subunits encoded by both nuclear and mtDNA genes. AD-related impairments in mitochondrial complex I in the brain and platelets have been observed^84^. These findings in our AA sample overlap with evidence from many studies for mitochondrial energy pathway involvement in AD^85^ and are supported by results from an AD DGE study in a large EA sample^16^. Sekar et al. examined posterior cingulate astrocytes extracted from 10 AD cases and 10 controls using laser microdissection and found lower expression in AD cases of several mitochondrial genes including *NDUFA4L2*^86^, which was nominally downregulated in our study (p=0.03). Multiple groups have identified mitochondrial dysfunction as an important cause of increased oxidative stress in brain tissue and suggest that this pathway is a potential druggable target for AD treatment and prevention^87–89^.

This study has several notable limitations. It should be acknowledged that this cohort of brain donors may not be representative of AA AD cases and controls more broadly. Although this is the largest study of post-mortem brain tissue from AA AD cases and controls, the sample is much smaller than other DGE studies conducted in EA autopsy samples (e.g. ^26,27^). Additionally, there is substantial variability in the collection and availability of neuropathological data and antemortem information including cognitive test data and history of relevant conditions (e.g., cardiovascular and TIA events, diabetes, or other medical complications) that precluded incorporation of these factors in our analyses. Finally, because this study was performed using bulk RNA sequencing data, we could not consider cell type specificity of our findings or detect associations evident only in underrepresented cell types. Subsequent studies could overcome this limitation through single-cell sequencing technology^90^ or deconvolution methods^91^.

In summary, we identified many genes that are differentially expressed in PFC tissue obtained from AA AD cases and controls. Several of the most significant DEGs were observed in EA brain donors, most notably *ADAMTS2*, whereas other novel DEGs and eQTLs may have been identified in AA individuals only due to population differences in the frequency of genetic variants or other risk factors. Our findings support the hypothesis that reelin and poor bioenergetics due to mitochondrial dysfunction play a role in AD susceptibility, and may lead to new hypotheses about pathogenic mechanisms, targets for drug development, and biomarkers for risk assessment and profiling subjects for clinical trials. This study also reinforces the value of diversity in genomic studies.

## Acknowledgements

We would like to acknowledge the contributions of Dr. Rachel Whitmer, Dr. Charles DeCarli, Erin E. Franklin, and Dr. Oscar Lopez.

## Data Availability

In accordance with National Institute of Aging (NIA) funding policies, the data underlying this paper will be deposited in the National Institute on Aging Genetics of Alzheimer’s Disease Data Storage Site (NIAGADS; https://www.niagads.org) after final study publication.

