## Supplementary Materials for "Transcriptome-wide association study of Alzheimer disease reveals many differentially expressed genes and multiple biological pathways in brain tissue from African American donors"

### Supplementary Materials for “A gene expression study of Alzheimer’s disease in post-mortem tissue from African American donors.” by M.W. Logue et al.

**Supplementary Figure 1:** Boxplot of the 6 most significant differentially expressed genes.

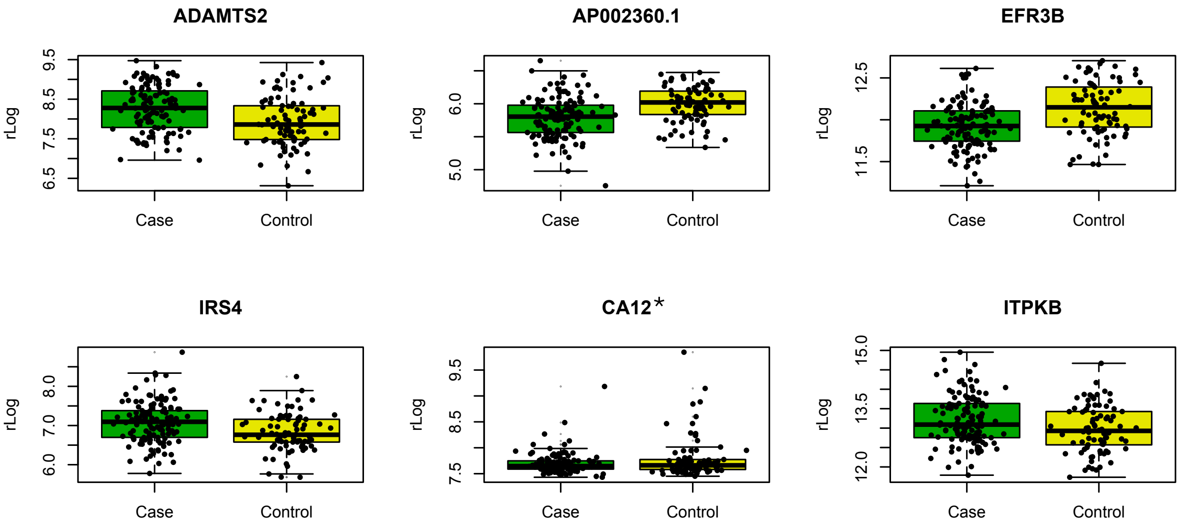


* Excludes an outlier with high expression in controls to preserve detail.

**Supplementary Figure 2:** Correlation of effect sizes from the sensitivity analyses for adjustment for PMI, and B) the exclusion of young (age <60) donors from the analyses. The effect sizes -$\log_{10}$ P-values were highly correlated for the PMI sensitivity analysis (A and B) and for the age sensitivity analysis (C and D). The correlations were all very high for the effect sizes and $\log_{10}$ P-values (r>0.90).

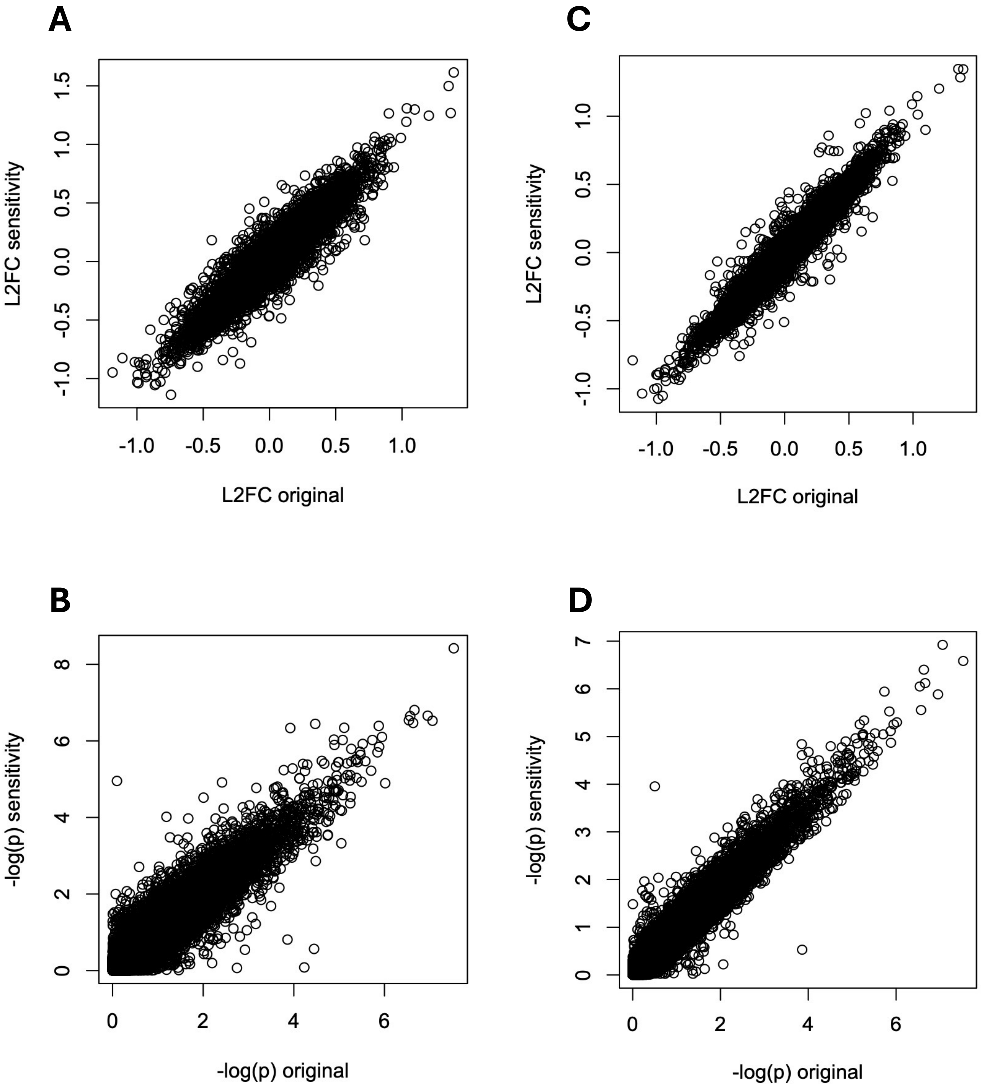

**Supplementary Table 1: SEE ATTACHMENT.** Table of the 482 transcriptome-wide significant associations between gene expression and AD case status.

**Supplementary Table 2:** Results of sensitivity analyses examining the impact of PMI and excluding controls with age at death < 60 for the top 20 differentially expressed genes.

| | PMI Sensitivity Analysis | | | Age Sensitivity Analysis | | |
|---|---|---|---|---|---|---|
| Gene | L2FC | P | Padj | L2FC | P | Padj |
| ADAMTS2 | 0.65 | 3.81E-09 | 0.00013 | 0.56 | 2.60E-07 | 0.0044 |
| AP002360.1 | -0.37 | 2.98E-07 | 0.0014 | -0.36 | 1.19E-07 | 0.0040 |
| EFR3B | -0.26 | 2.20E-07 | 0.0014 | -0.24 | 1.31E-06 | 0.0063 |
| IRS4 | 0.51 | 1.57E-07 | 0.0014 | 0.46 | 7.57E-07 | 0.0060 |
| CA12 | -0.83 | 3.40E-07 | 0.0014 | -0.79 | 3.96E-07 | 0.0044 |
| ITPKB | 0.34 | 2.25E-07 | 0.0014 | 0.29 | 2.78E-06 | 0.011 |
| PDE10A | -0.50 | 2.89E-07 | 0.0014 | -0.46 | 8.91E-07 | 0.0060 |
| TDRKH | -0.16 | 1.27E-05 | 0.0079 | -0.16 | 5.02E-06 | 0.014 |
| LINC01182 | -0.71 | 7.89E-07 | 0.0021 | -0.63 | 5.57E-06 | 0.014 |
| TFCP2 | -0.17 | 1.43E-06 | 0.0027 | -0.15 | 1.35E-05 | 0.019 |
| AL158071.3 | 0.58 | 4.07E-07 | 0.0014 | 0.49 | 7.68E-06 | 0.017 |
| EHHADH | -0.32 | 1.99E-06 | 0.0032 | -0.30 | 2.98E-06 | 0.011 |
| PIEZO2 | 0.45 | 9.49E-07 | 0.0021 | 0.42 | 1.14E-06 | 0.0063 |
| P2RY1 | -0.44 | 6.02E-06 | 0.0053 | -0.41 | 9.00E-06 | 0.017 |
| AC018647.2 | -0.26 | 3.39E-06 | 0.0040 | -0.23 | 2.02E-05 | 0.021 |
| NOX4 | -0.45 | 1.20E-06 | 0.0025 | -0.41 | 8.57E-06 | 0.017 |
| ECHS1 | -0.17 | 1.17E-05 | 0.0077 | -0.16 | 2.67E-05 | 0.022 |
| ZCCHC14 | 0.15 | 2.32E-06 | 0.0035 | 0.12 | 3.88E-05 | 0.025 |
| LINC01094 | 0.46 | 1.92E-06 | 0.0032 | 0.40 | 1.59E-05 | 0.019 |
| AP001180.1 | 1.50 | 3.58E-06 | 0.0040 | 1.35 | 1.06E-05 | 0.017 |

**Supplementary Table 3:** Tests for differential gene expression of genome-wide significant loci from AD GWAS in EA (Bellenguez et al. 2022) and AA (Sherva et al. 2023) cohorts, and other genes implicated in prior AA GWAS or candidate gene association studies (Logue et al. 2023). False discovery rate multiple testing corrections performed sequentially within each category.

| EA AD GWAS | | | | | | | | AA AD GWAS | | | |
|---|---|---|---|---|---|---|---|---|---|---|---|
| **Gene** | **L2FC** | **P** | **$p_{adj}$** | **Gene** | **L2FC** | **P** | **$p_{adj}$** | **Gene** | **L2FC** | **P** | **$p_{adj}$** |
| **WNT3** | 0.16 | 0.052 | 0.0017 | DOC2A | 0.048 | 0.068 | 0.48 | **KIF17** | -0.17 | 0.021 | 0.17 |
| **ICA1** | -0.17 | 0.068 | 0.010 | IL34 | 0.061 | 0.091 | 0.50 | **SH2D5** | -0.16 | 0.035 | 0.17 |
| **PLEKHA1** | 0.11 | 0.041 | 0.010 | ADAMTS1 | 0.072 | 0.11 | 0.51 | **DCBLD1** | -0.093 | 0.043 | 0.17 |
| **FERMT2** | -0.11 | 0.045 | 0.011 | BIN1 | -0.029 | 0.045 | 0.52 | **POLR2E** | -0.086 | 0.052 | 0.17 |
| **APOE** | -0.20 | 0.079 | 0.012 | CR1 | 0.086 | 0.14 | 0.53 | **NUS1** | 0.060 | 0.16 | 0.42 |
| **SORL1** | -0.10 | 0.042 | 0.014 | SIGLEC11 | 0.095 | 0.16 | 0.54 | **EIF4G3** | -0.028 | 0.27 | 0.53 |
| **PRDM7** | 0.55 | 0.232 | 0.017 | FOXF1 | -0.061 | 0.10 | 0.55 | **TREML1** | -0.15 | 0.29 | 0.53 |
| **SNX1** | -0.069 | 0.029 | 0.017 | SLC24A4 | 0.037 | 0.061 | 0.55 | **GOPC** | -0.038 | 0.33 | 0.54 |
| **SLC2A4RG** | -0.15 | 0.063 | 0.019 | RHOH | 0.11 | 0.19 | 0.55 | **ARHGAP45** | 0.035 | 0.63 | 0.91 |
| **CLNK** | 0.68 | 0.30 | 0.022 | JAZF1 | -0.024 | 0.040 | 0.56 | **TMCO4** | -0.017 | 0.82 | 0.97 |
| **HS3ST5** | -0.14 | 0.068 | 0.036 | CD2AP | 0.022 | 0.039 | 0.57 | **CDA** | -0.029 | 0.85 | 0.97 |
| **COX7C** | -0.10 | 0.052 | 0.047 | WDR81 | -0.029 | 0.052 | 0.57 | **CNN2** | -0.009 | 0.93 | 0.97 |
| **TSPOAP1** | -0.11 | 0.054 | 0.050 | TMEM106B | -0.030 | 0.054 | 0.58 | **ROS1** | 0.005 | 0.97 | 0.97 |
| **TPCN1** | -0.11 | 0.059 | 0.066 | GRN | -0.034 | 0.061 | 0.58 | **ANTXR2** | 0.13 | 0.021 | 0.24 |
| **CTSB** | -0.066 | 0.036 | 0.067 | CASS4 | 0.055 | 0.11 | 0.60 | **GRIN3B** | -0.34 | 0.078 | 0.32 |
| **BCKDK** | -0.083 | 0.046 | 0.069 | LILRB2 | -0.076 | 0.15 | 0.61 | **EML6** | -0.069 | 0.087 | 0.32 |
| **TREM2** | 0.21 | 0.12 | 0.074 | CLU | 0.023 | 0.044 | 0.61 | **EFHB** | -0.27 | 0.18 | 0.47 |
| **SORT1** | 0.064 | 0.036 | 0.075 | SPI1 | -0.038 | 0.075 | 0.61 | **TRPS1** | -0.042 | 0.21 | 0.47 |
| **RBCK1** | -0.089 | 0.051 | 0.080 | EPDR1 | -0.033 | 0.067 | 0.63 | **FHOD3** | 0.037 | 0.35 | 0.65 |
| **TSPAN14** | 0.079 | 0.046 | 0.08 | ABCA7 | 0.038 | 0.080 | 0.63 | **ROBO1** | -0.026 | 0.46 | 0.68 |
| **MS4A4A** | 0.20 | 0.14 | 0.14 | ACE | -0.040 | 0.086 | 0.64 | **AL355353.1** | -0.27 | 0.49 | 0.68 |
| **MAF** | 0.089 | 0.062 | 0.15 | HLA-DQA1 | 0.075 | 0.19 | 0.69 | **ITGA11** | 0.022 | 0.75 | 0.90 |
| **SPDYE3** | 0.091 | 0.065 | 0.16 | ABI3 | 0.036 | 0.093 | 0.70 | **KCNH8** | -0.019 | 0.83 | 0.90 |
| **PRKD3** | 0.043 | 0.031 | 0.18 | KLF16 | 0.024 | 0.063 | 0.70 | **EGFLAM** | -0.020 | 0.90 | 0.90 |
| **APH1B** | -0.055 | 0.041 | 0.18 | PLCG2 | 0.024 | 0.067 | 0.72 | | | | |
| **BLNK** | -0.11 | 0.086 | 0.19 | EED | -0.011 | 0.036 | 0.75 | | | | |

| | | | | | | | | | | | |
|---|---|---|---|---|---|---|---|---|---|---|---|
| **ABCA1** | 0.083 | 0.063 | 0.19 | TREML2 | -0.072 | 0.23 | 0.76 | **Other AA AD Genetics Studies** | | | |
| **INPP5D** | 0.082 | 0.072 | 0.25 | MINDY2 | -0.013 | 0.041 | 0.76 | **IGF1R** | 0.11 | 0.0013 | 0.019 |
| **RASGEF1C** | -0.092 | 0.080 | 0.25 | SPPL2A | 0.013 | 0.043 | 0.76 | **COBL** | 0.079 | 0.083 | 0.31 |
| **PTK2B** | -0.058 | 0.051 | 0.26 | TNIP1 | 0.011 | 0.038 | 0.77 | **AKAP9** | 0.070 | 0.083 | 0.31 |
| **SEC61G** | -0.059 | 0.056 | 0.29 | SHARPIN | -0.014 | 0.054 | 0.79 | **PON2** | -0.075 | 0.11 | 0.31 |
| **APP** | -0.030 | 0.031 | 0.34 | CTSH | 0.013 | 0.055 | 0.81 | **NOS3** | -0.17 | 0.11 | 0.31 |
| **UNC5CL** | -0.093 | 0.10 | 0.35 | EPHA1 | 0.026 | 0.15 | 0.86 | **PSEN1** | -0.044 | 0.12 | 0.31 |
| **MYO15A** | 0.067 | 0.071 | 0.35 | ADAM17 | -0.0053 | 0.031 | 0.87 | **PSEN2** | -0.056 | 0.18 | 0.34 |
| **ANKH** | 0.023 | 0.027 | 0.40 | SCIMP | -0.0073 | 0.062 | 0.91 | **PICALM** | -0.052 | 0.18 | 0.34 |
| **NCK2** | 0.031 | 0.038 | 0.41 | IDUA | -0.0058 | 0.069 | 0.93 | **SORCS1** | -0.072 | 0.25 | 0.41 |
| **ANK3** | -0.022 | 0.028 | 0.44 | MME | -0.026 | 0.43 | 0.95 | **SYNPR** | 0.068 | 0.54 | 0.80 |
| **USP6NL** | -0.027 | 0.036 | 0.45 | WDR12 | -0.00040 | 0.036 | 0.99 | **SELP** | 0.091 | 0.65 | 0.81 |
| **UMAD1** | -0.032 | 0.043 | 0.46 | | | | | **ARRDC4** | -0.040 | 0.69 | 0.81 |
| | | | | | | | | **F5** | -0.039 | 0.71 | 0.81 |
| | | | | | | | | **TTR** | -0.094 | 0.90 | 0.94 |
| | | | | | | | | **DIO2** | -0.24 | 0.94 | 0.94 |

**Supplementary Table 4:** GOseq analysis of the differentially expressed genes and of the members of the WGCNA networks. Members of the GO category or network that are differentially expressed at the $p<0.05$ level are labeled + and - to indicate the direction of effect, while ++ and -- indicate that the gene is associated at the transcriptome-wide ($p_{adj}<0.05$) level. The top 10 enriched categories are listed for gene sets with significant enrichments. For gene categories with no corrected significant GO terms, the top 5 GO terms are listed.

| network | ontology | category | term | Num DE in category | Num in category | p | padj | genes |
|---|---|---|---|---|---|---|---|---|
| **DE Genes** | BP | GO:0055114 | oxidation-reduction process | 55 | 927 | 6.38E-11 | 3.07E-07 | ABCD2--,ACAA2--,ADHFE1--,ADIPOR1--,ALDH7A1--,ALDH9A1--,BCKDHB--,BPGM--,CCS--,COX15--,COX4I1--,COX6B1--,CPT2--,CYP2J2--,DDO--,DECR1--,DHFR2--,ECHS1--,EGLN3--,EHHADH--,ETFA--,ETFB--,F8--,FAXDC2--,FDX2--,GLUD1--,HADHA--,HSD11B2++,HSD17B7++,IDH2--,IRS2++,MSRB1--,NDUFA1--,NDUFA2--,NDUFA3--,NDUFA6--,NDUFA7--,NDUFB1--,NDUFB2--,NDUFB3--,NDUFB8--,NDUFS3--,NDUFS5--,NOX4--,PANK2++,PFKP++,PLA2G7--,PRDX2--,PRDX5--,STEAP3++,STEAP4++,TM7SF2--,TXN2--,TXNDC12--,UQCRC2-- |
| | BP | GO:0044281 | small molecule metabolic process | 86 | 1945 | 7.72E-11 | 3.07E-07 | ABCC2++,ABCD2--,ACAA2--,ADA2--,ADAL--,ADHFE1--,ADIPOR1--,AGPAT1--,AIG1--,ALDH7A1--,ARV1--,ATP5F1B--,ATP5F1C--,ATP5MF--,ATP5MG--,ATP5PB--,BCKDHB--,BPGM--,CD244--,CHST6++,COQ5--,COX15--,COX4I1--,COX6B1--,CPT2--,CYP2J2--,DDO--,DECR1--,DHFR2--,ECHS1--,EGLN3--,EHHADH--,ENOPH1--,ERG28--,ETFA--,ETFB--,FDX2--,GLUD1--,HADHA--,HDAC4++,HDDC3--,HKDC1--,HSD17B7++,IDH2--,IRS2++,ITPKB++,LARS2--,LCLAT1--,MCCC2--,MPC1--,NDST3--,NDUFA1--,NDUFA2--,NDUFA3--,NDUFA6--,NDUFA7--,NDUFB1--,NDUFB2--,NDUFB3--,NDUFB8--,NDUFS3--,NDUFS5--,NFS1--,NOX4--,NUDT7--,P2RY1--,PANK2++,PDE10A--,PDE7B--,PDZD11--,PFKP++,PLCE1++,PPT1--,PRKAG1--,PRTFDC1--,PSMC3--,PSME3--,RIDA--,SDSL--,SEC13--,SLC7A2++,SRR--,TM7SF2--,TXN2--,UQCRC2--,VCAN++ |
| | MF | GO:0016491 | oxidoreductase activity | 45 | 686 | 8.61E-10 | 2.28E-06 | ADHFE1--,ALDH7A1--,ALDH9A1--,BCKDHB--,CCS--,COX15--,COX4I1--,COX6B1--,CYP2J2--,DDO--,DECR1--,DHFR2--,EGLN3--,EHHADH--,ETFA--,ETFB--,F8--,FAXDC2--,FDX2--,GLUD1--,HADHA--,HBA1--,HSD11B2++,HSD17B7++,IDH2--,MSRB1--,NDUFA1--,NDUFA2--,NDUFA3--,NDUFA6--,NDUFA7--,NDUFB1--,NDUFB2--,NDUFB3--,NDUFB8--,NDUFS3--,NDUFS5--,NOX4--,PRDX2--,PRDX5--,STEAP3++,STEAP4++,TM7SF2--,TXN2--,TXNDC12-- |
| | BP | GO:0072521 | purine-containing compound metabolic process | 39 | 565 | 1.64E-09 | 3.26E-06 | ADA2--,ADAL--,ATP5F1B--,ATP5F1C--,ATP5MF--,ATP5MG--,ATP5PB--,BPGM--,COX15--,COX4I1--,COX6B1--,ENOPH1--,HDAC4++,HDDC3--,HKDC1--,MCCC2--,MPC1--,NDUFA1--,NDUFA2--,NDUFA3--,NDUFA6--,NDUFA7--,NDUFB1--,NDUFB2--,NDUFB3--,NDUFB8--,NDUFS3--,NDUFS5--,NFS1--,NUDT7--,PANK2++,PDE10A--,PDE7B--,PFKP++,PPT1--,PRKAG1--,PRTFDC1--,SEC13--,UQCRC2-- |
| | BP | GO:0006119 | oxidative phosphorylation | 20 | 128 | 3.15E-09 | 5.01E-06 | ATP5F1B--,ATP5F1C--,ATP5MF--,ATP5MG--,ATP5PB--,COX15--,COX4I1--,COX6B1--,NDUFA1--,NDUFA2--,NDUFA3--,NDUFA6--,NDUFA7--,NDUFB1--,NDUFB2--,NDUFB3--,NDUFB8--,NDUFS3--,NDUFS5--,UQCRC2-- |
| | BP | GO:0006091 | generation of precursor metabolites and energy | 35 | 485 | 4.68E-09 | 6.19E-06 | ATP5F1B--,ATP5F1C--,ATP5MF--,ATP5MG--,ATP5PB--,BPGM--,COX15--,COX4I1--,COX6B1--,ETFA--,ETFB--,FDX2--,HDAC4++,HKDC1--,IDH2--,IRS2++,NDUFA1--,NDUFA2--,NDUFA3--,NDUFA6--,NDUFA7--,NDUFB1--,NDUFB2--,NDUFB3--,NDUFB8--,NDUFS3--,NDUFS5--,NOX4--,PANK2++,PFKP++,PRKAG1--,SEC13--,STEAP4++,UQCRC2--,VGF-- |
| | BP | GO:0017144 | drug metabolic process | 45 | 771 | 9.53E-09 | 1.08E-05 | ABCC2++,ADA2--,ADAL--,ATP5F1B--,ATP5F1C--,ATP5MF--,ATP5MG--,ATP5PB--,BPGM--,COX15--,COX4I1--,COX6B1--,CYP2J2--,DHFR2--,ENOPH1--,HBA1--,HDAC4++,HKDC1--,IDH2--,MCCC2--,NDUFA1--,NDUFA2--,NDUFA3--,NDUFA6--,NDUFA7--,NDUFB1--,NDUFB2--,NDUFB3--,NDUFB8--,NDUFS3--,NDUFS5--,PDZD11-- |

| | | | | | | | | |
|---|---|---|---|---|---|---|---|---|
| | | | | | | | | ,PFKP++,PLA2G7--,PRDX2--,PRDX5--,PRKAG1--,RIDA--,SDSL--,SEC13--,SLC7A2++,SRR--,TGFB2++,UQCRC2--,VCAN++ |
| | BP | GO:0046034 | ATP metabolic process | 26 | 282 | 1.43E-08 | 1.36E-05 | ATP5F1B--,ATP5F1C--,ATP5MF--,ATP5MG--,ATP5PB--,BPGM--,COX15--,COX4I1--,COX6B1--,HDAC4++,HKDC1--,NDUFA1--,NDUFA2--,NDUFA3--,NDUFA6--,NDUFA7--,NDUFB1--,NDUFB2--,NDUFB3--,NDUFB8--,NDUFS3--,NDUFS5--,PFKP++,PRKAG1--,SEC13--,UQCRC2-- |
| | BP | GO:0009150 | purine ribonucleotide metabolic process | 35 | 506 | 1.54E-08 | 1.36E-05 | ATP5F1B--,ATP5F1C--,ATP5MF--,ATP5MG--,ATP5PB--,BPGM--,COX15--,COX4I1--,COX6B1--,HDAC4++,HDDC3--,HKDC1--,MCCC2--,MPC1--,NDUFA1--,NDUFA2--,NDUFA3--,NDUFA6--,NDUFA7--,NDUFB1--,NDUFB2--,NDUFB3--,NDUFB8--,NDUFS3--,NDUFS5--,NFS1--,NUDT7--,PANK2++,PDE10A--,PDE7B--,PFKP++,PPT1--,PRKAG1--,SEC13--,UQCRC2-- |
| | BP | GO:0009205 | purine ribonucleoside triphosphate metabolic process | 27 | 311 | 2.62E-08 | 1.99E-05 | ATP5F1B--,ATP5F1C--,ATP5MF--,ATP5MG--,ATP5PB--,BPGM--,COX15--,COX4I1--,COX6B1--,HDAC4++,HKDC1--,NDUFA1--,NDUFA2--,NDUFA3--,NDUFA6--,NDUFA7--,NDUFB1--,NDUFB2--,NDUFB3--,NDUFB8--,NDUFS3--,NDUFS5--,NFS1--,PFKP++,PRKAG1--,SEC13--,UQCRC2-- |
| **blue** | MF | GO:0003676 | nucleic acid binding | 800 | 3378 | 9.33E-39 | 6.85E-35 | AHCTF1+,AIMP1-,ATF6-,ATR-,ATRX+,BLM+,BLZF1-,CAPRIN1-,CDK13+,CHD1+,CLNS1A-,CLOCK+,CPSF3-,CREBRF+,CRLF3+,DCAF13-,DDX50-,DEK+,DNAJC2+,EIF2A-,EIF2B1-,EIF2S1-,EIF2S3-,EIF3M-,EIF4E-,EPRS1-,ERH-,ETV1-,FASTKD3-,FIP1L1-,FOXO3+,G3BP1+,GDI2-,GFM1-,GNL2-,GTF2B-,GTF2H5-,H4C3-,HMGN1-,LCOR++,LCORL+,LIG4-,LIN9+,LSM3-,MAGOH-,MAK16-,MBD2+,MEIS2-,MFAP1-,MIER3+,MLF1-,MORC3+,MRPL13-,MRPS18C-,MRPS28-,MRPS35-,MRPS9--,NFAT5+,NFE2L3+,NUDT4+,NUP42-,ORC3-,OTUD4+,PA2G4-,PDIA3-,PIN4-,POLI-,POLR2K--,POU5F2+,PRKDC-,PRMT3-,PRPF38B+,PRPF40A+,PRPF4B+,PURB+,PUS7+,RBBP6+,RBM48+,RBMXL1-,RC3H2+,REL+,RORA+,RPF1-,SAP18-,SETDB2+,SF3B6-,SIRT1+,SLIRP-,SMAD4+,SMARCA1++,SNRPD1-,SNRPE-,SNW1-,SREK1+,SRP19+,SRP54-,SRP9-,SRSF10+,SUCLG1-,SUPT3H-,TBCA-,TDRD3-,TENT4B+,THOC2+,THOC7-,TIPIN-,TOPORS+,TSN-,TUT4+,WDHD1--,YWHAE-,YY1+,ZBTB1+,ZBTB44+,ZC3H13-,ZCCHC7-,ZCCHC8+,ZCCHC9-,ZFC3H1+,ZFP3-,ZMYM4-,ZNF141+,ZNF264+,ZNF280C-,ZNF292++,ZNF345+,ZNF383+,ZNF431+,ZNF441-,ZNF442-,ZNF484+,ZNF502-,ZNF546+,ZNF585A-,ZNF627--,ZNF639+,ZNF737-,ZRANB3- |
| **blue** | BP | GO:0090304 | nucleic acid metabolic process | 936 | 4319 | 2.66E-30 | 9.76E-27 | ADAT2-,AHCTF1+,AIMP1-,ANKRD31-,ARMCX3-,ASF1A-,ATF6-,ATR-,ATRX+,BLM+,BLZF1-,BMI1+,BMT2+,BRWD1+,CBX5+,CDK13+,CETN2-,CHD1+,CHURC1--,CLNS1A-,CLOCK+,CNOT10--,CPSF3-,CREBRF+,CREG1-,CRLF3+,CTH-,DCAF13-,DEK+,DNAJC2+,DUS4L-,ECD-,EGLN1+,EIF2A-,ELOC-,EPC1+,EPC2++,EPRS1-,ETV1-,FANCF-,FASTKD3-,FIP1L1-,FOXO3+,G3BP1+,GALR1-,GEMIN2-,GTF2A2-,GTF2B-,GTF2E1-,GTF2H3-,GTF2H5-,H4C3-,HIGD1A-,HMGN1-,IK-,INO80D+,INTS6+,JAK2+,LCOR++,LCORL+,LIG4-,LIN9+,LSM3-,MAGOH-,MAP3K2+,MBD2+,MED4-,MEIS2-,METTL15-,MFAP1-,MIER3+,MLF1-,MRPS9--,NAA16++,NBDY-,NCOA4-,NFAT5+,NFE2L3+,ORC3-,OTUD4+,PA2G4-,PHF3+,PLRG1-,POLI-,POLR2K--,POU5F2+,PPP4R2+,PRDX3-,PRKDC-,PRMT3-,PRMT6-,PRPF38B+,PRPF40A+,PRPF4B+,PSMD10-,PSMD14-,PURB+,PUS7+,QTRT2-,RAC1-,RBBP6+,RBMXL1-,RC3H2+,REL+,RESF1+,RHOQ+,RIOK1-,RNF14-,RNF168+,RORA+,RPF1-,SAP18-,SCARNA7+,SETD6+,SETDB2+,SF3B6-,SIRT1+,SLIRP-,SMAD4+,SMARCA1++,SMC5+,SNRPD1-,SNRPE-,SNW1-,SREK1+,SRSF10+,SUPT3H-,TATDN1-,TATDN3-,TDRD3-,TENT2-,TENT4B+,THOC2+,THOC7-,TIPIN-,TOPORS+,TRIP12-,TSN-,TTC21B+,TUT4+,UFL1-,WDHD1--,XRCC4-,YY1+,ZBTB1+,ZBTB44+,ZC3H12C+,ZC3H13-,ZCCHC8+,ZFC3H1+,ZFP3-,ZMYM4-,ZNF141+,ZNF264+,ZNF280C-,ZNF292++,ZNF345+,ZNF383+,ZNF431+,ZNF441-,ZNF442-,ZNF484+,ZNF502-,ZNF546+,ZNF585A-,ZNF627--,ZNF639+,ZNF737-,ZRANB3- |
| **blue** | MF | GO:0003723 | RNA binding | 406 | 1537 | 2.55E-29 | 6.25E-26 | AIMP1-,CAPRIN1-,CDK13+,CLNS1A-,CPSF3-,DCAF13-,DDX50-,DEK+,DNAJC2+,EIF2A-,EIF2B1-,EIF2S1-,EIF2S3-,EIF3M-,EIF4E-,EPRS1-,ERH-,FASTKD3-,FIP1L1-,G3BP1+,GDI2-,GFM1-,GNL2-,H4C3-,LSM3-,MAGOH-,MAK16-,MBD2+,MFAP1-,MORC3+,MRPL13-,MRPS18C-,MRPS28-,MRPS35-,MRPS9--,NUDT4+,NUP42-,OTUD4+,PA2G4-,PDIA3- |

| | | | | | | | | |
|---|---|---|---|---|---|---|---|---|
| | | | | | | | | ,PIN4-,PRKDC-,PRPF38B+,PRPF40A+,PRPF4B+,PURB+,PUS7+,RBBP6+,RBM48+,RBMXL1-,RC3H2+,RPF1-,SAP18-,SF3B6-,SLIRP-,SNRPD1-,SNRPE-,SNW1-,SREK1+,SRP19+,SRP54-,SRP9-,SRSF10+,SUCLG1-,TBCA-,TDRD3-,TENT4B+,THOC2+,THOC7-,TSN-,TUT4+,YWHAE-,YY1+,ZC3H13-,ZCCHC7-,ZCCHC8+,ZCCHC9-,ZFC3H1+ |
| **blue** | BP | GO:0034641 | cellular nitrogen compound metabolic process | 1117 | 5456 | 1.75E-28 | 3.21E-25 | ADAL--,ADAT2-,AFG1L-,AHCTF1+,AIMP1-,ANKRD31-,ARMCX3-,ASF1A-,ATF6-,ATIC-,ATP5MG--,ATP5PB--,ATR-,ATRX+,BLM+,BLZF1-,BMI1+,BMT2+,BPNT1-,BRWD1+,CAPRIN1-,CBX5+,CDK13+,CETN2-,CHD1+,CHURC1--,CLNS1A-,CLOCK+,CLPX-,CNOT10--,CPSF3-,CREBRF+,CREG1-,CRLF3+,CTH-,DCAF13-,DDAH1-,DEK+,DNAJC2+,DUS4L-,ECD-,EGLN1+,EIF2A-,EIF2B1-,EIF2S1-,EIF2S3-,EIF3M-,EIF4E-,ELOC-,EPC1+,EPC2++,EPRS1-,ERH-,ETV1-,FANCF-,FASTKD3-,FIP1L1-,FOXO3+,G3BP1+,GALR1-,GART-,GEMIN2-,GFM1-,GLO1-,GNPNAT1+,GPD2-,GTF2A2-,GTF2B-,GTF2E1-,GTF2H3-,GTF2H5-,H4C3-,HIGD1A-,HMGN1-,HSD17B12-,IK-,INO80D+,INTS6+,ITGB8+,JAK2+,LCOR++,LCORL+,LIAS-,LIG4-,LIN9+,LRRK2-,LSM3-,MAGOH-,MAP3K2+,MBD2+,ME1-,ME2-,MED4-,MEIS2-,METTL15-,MFAP1-,MIER3+,MLF1-,MPC1--,MRPL13-,MRPL47-,MRPL51-,MRPS18C-,MRPS28-,MRPS35-,MRPS9--,NAA16++,NBDY-,NCOA4-,NDUFA12-,NDUFC1-,NDUFS4-,NFAT5+,NFE2L3+,NIPSNAP2-,NUDT12-,NUDT4+,NUP42-,NUP54-,ORC3-,OTUD4+,PA2G4-,PDHB-,PDHX-,PHF3+,PLRG1-,POLI-,POLR2K--,POU5F2+,PPP4R2+,PRDX3-,PRKDC-,PRMT3-,PRMT6-,PRPF38B+,PRPF40A+,PRPF4B+,PSMD10-,PSMD14-,PURB+,PUS7+,QTRT2-,RAC1-,RBBP6+,RBMXL1-,RC3H2+,REL+,RESF1+,RHOQ+,RIOK1-,RNF14-,RNF168+,RORA+,RPF1-,SAP18-,SCARNA7+,SDHD-,SETD6+,SETDB2+,SF3B6-,SIRT1+,SLC35A3+,SLIRP-,SMAD4+,SMARCA1++,SMC5+,SNRPD1-,SNRPE-,SNW1-,SPCS2-,SREK1+,SRP9-,SRSF10+,SUCLG1-,SUPT3H-,TATDN1-,TATDN3-,TDRD3-,TENT2-,TENT4B+,THOC2+,THOC7-,TIPIN-,TMA7-,TOPORS+,TRIP12-,TSN-,TTC21B+,TUT4+,UFL1-,WDHD1--,XRCC4-,YY1+,ZBTB1+,ZBTB44+,ZC3H12C+,ZC3H13-,ZCCHC8+,ZFC3H1+,ZFP3-,ZMYM4-,ZNF141+,ZNF264+,ZNF280C-,ZNF292++,ZNF345+,ZNF383+,ZNF431+,ZNF441-,ZNF442-,ZNF484+,ZNF502-,ZNF546+,ZNF585A-,ZNF627--,ZNF639+,ZNF737-,ZRANB3- |
| **blue** | BP | GO:0006139 | nucleobase-containing compound metabolic process | 1011 | 4885 | 6.93E-26 | 1.02E-22 | ADAL--,ADAT2-,AFG1L-,AHCTF1+,AIMP1-,ANKRD31-,ARMCX3-,ASF1A-,ATF6-,ATIC-,ATP5MG--,ATP5PB--,ATR-,ATRX+,BLM+,BLZF1-,BMI1+,BMT2+,BPNT1-,BRWD1+,CBX5+,CDK13+,CETN2-,CHD1+,CHURC1--,CLNS1A-,CLOCK+,CLPX-,CNOT10--,CPSF3-,CREBRF+,CREG1-,CRLF3+,CTH-,DCAF13-,DEK+,DNAJC2+,DUS4L-,ECD-,EGLN1+,EIF2A-,ELOC-,EPC1+,EPC2++,EPRS1-,ERH-,ETV1-,FANCF-,FASTKD3-,FIP1L1-,FOXO3+,G3BP1+,GALR1-,GART-,GEMIN2-,GNPNAT1+,GPD2-,GTF2A2-,GTF2B-,GTF2E1-,GTF2H3-,GTF2H5-,H4C3-,HIGD1A-,HMGN1-,HSD17B12-,IK-,INO80D+,INTS6+,JAK2+,LCOR++,LCORL+,LIG4-,LIN9+,LRRK2-,LSM3-,MAGOH-,MAP3K2+,MBD2+,ME1-,ME2-,MED4-,MEIS2-,METTL15-,MFAP1-,MIER3+,MLF1-,MPC1--,MRPS9--,NAA16++,NBDY-,NCOA4-,NDUFA12-,NDUFC1-,NDUFS4-,NFAT5+,NFE2L3+,NIPSNAP2-,NUDT12-,NUDT4+,NUP42-,NUP54-,ORC3-,OTUD4+,PA2G4-,PDHB-,PDHX-,PHF3+,PLRG1-,POLI-,POLR2K--,POU5F2+,PPP4R2+,PRDX3-,PRKDC-,PRMT3-,PRMT6-,PRPF38B+,PRPF40A+,PRPF4B+,PSMD10-,PSMD14-,PURB+,PUS7+,QTRT2-,RAC1-,RBBP6+,RBMXL1-,RC3H2+,REL+,RESF1+,RHOQ+,RIOK1-,RNF14-,RNF168+,RORA+,RPF1-,SAP18-,SCARNA7+,SDHD-,SETD6+,SETDB2+,SF3B6-,SIRT1+,SLC35A3+,SLIRP-,SMAD4+,SMARCA1++,SMC5+,SNRPD1-,SNRPE-,SNW1-,SREK1+,SRSF10+,SUCLG1-,SUPT3H-,TATDN1-,TATDN3-,TDRD3-,TENT2-,TENT4B+,THOC2+,THOC7-,TIPIN-,TOPORS+,TRIP12-,TSN-,TTC21B+,TUT4+,UFL1-,WDHD1--,XRCC4-,YY1+,ZBTB1+,ZBTB44+,ZC3H12C+,ZC3H13-,ZCCHC8+,ZFC3H1+,ZFP3-,ZMYM4-,ZNF141+,ZNF264+,ZNF280C-,ZNF292++,ZNF345+,ZNF383+,ZNF431+,ZNF441-,ZNF442-,ZNF484+,ZNF502-,ZNF546+,ZNF585A-,ZNF627--,ZNF639+,ZNF737-,ZRANB3- |

| | | | | | | | | |
|---|---|---|---|---|---|---|---|---|
| **blue** | BP | GO:0006396 | RNA processing | 261 | 897 | 9.19E-26 | 1.12E-22 | ADAT2-,BMT2+,CDK13+,CLNS1A-,CPSF3-,DCAF13-,DUS4L-,ECD-,FIP1L1-,GEMIN2-,GTF2H3-,GTF2H5-,IK-,INTS6+,LSM3-,MAGOH-,METTL15-,MFAP1-,MRPS9--,NBDY-,PA2G4-,PLRG1-,POLR2K--,PPP4R2+,PRPF38B+,PRPF40A+,PRPF4B+,PUS7+,QTRT2-,RBBP6+,RBMXL1-,RIOK1-,RPF1-,SAP18-,SCARNA7+,SF3B6-,SIRT1+,SNRPD1-,SNRPE-,SNW1-,SREK1+,SRSF10+,TENT2-,TENT4B+,THOC2+,THOC7-,TSN-,TUT4+,ZC3H13-,ZCCHC8+,ZFC3H1+ |
| **blue** | BP | GO:0046483 | heterocycle metabolic process | 1025 | 4987 | 2.94E-25 | 3.08E-22 | ADAL--,ADAT2-,AFG1L-,AHCTF1+,AIMP1-,ANKRD31-,ARMCX3-,ASF1A-,ATF6-,ATIC-,ATP5MG--,ATP5PB--,ATR-,ATRX+,BLM+,BLZF1-,BMI1+,BMT2+,BPNT1-,BRWD1+,CBX5+,CDK13+,CETN2-,CHD1+,CHURC1--,CLNS1A-,CLOCK+,CLPX-,CNOT10--,CPSF3-,CREBRF+,CREG1-,CRLF3+,CTH-,DCAF13-,DEK+,DNAJC2+,DUS4L-,ECD-,EGLN1+,EIF2A-,ELOC-,EPC1+,EPC2++,EPRS1-,ERH-,ETV1-,FANCF-,FASTKD3-,FIP1L1-,FOXO3+,G3BP1+,GALR1-,GART-,GEMIN2-,GNPNAT1+,GPD2-,GTF2A2-,GTF2B-,GTF2E1-,GTF2H3-,GTF2H5-,H4C3-,HIGD1A-,HMGN1-,HSD17B12-,IK-,INO80D+,INTS6+,JAK2+,LCOR++,LCORL+,LIAS-,LIG4-,LIN9+,LRRK2-,LSM3-,MAGOH-,MAP3K2+,MBD2+,ME1-,ME2-,MED4-,MEIS2-,METTL15-,MFAP1-,MIER3+,MLF1-,MMUT-,MPC1--,MRPS9--,MTR-,NAA16++,NBDY-,NCOA4-,NDUFA12-,NDUFC1-,NDUFS4-,NFAT5+,NFE2L3+,NIPSNAP2-,NUDT12-,NUDT4+,NUP42-,NUP54-,ORC3-,OTUD4+,PA2G4-,PDHB-,PDHX-,PHF3+,PLRG1-,POLI-,POLR2K--,POU5F2+,PPP4R2+,PRDX3-,PRKDC-,PRMT3-,PRMT6-,PRPF38B+,PRPF40A+,PRPF4B+,PSMD10-,PSMD14-,PURB+,PUS7+,QTRT2-,RAC1-,RBBP6+,RBMXL1-,RC3H2+,REL+,RESF1+,RHOQ+,RIOK1-,RNF14-,RNF168+,RORA+,RPF1-,SAP18-,SCARNA7+,SDHD-,SETD6+,SETDB2+,SF3B6-,SIRT1+,SLC35A3+,SLIRP-,SMAD4+,SMARCA1++,SMC5+,SNRPD1-,SNRPE-,SNW1-,SREK1+,SRSF10+,SUCLG1-,SUPT3H-,TATDN1-,TATDN3-,TDRD3-,TENT2-,TENT4B+,THOC2+,THOC7-,TIPIN-,TOPORS+,TRIP12-,TSN-,TTC21B+,TUT4+,UFL1-,WDHD1--,XRCC4-,YY1+,ZBTB1+,ZBTB44+,ZC3H12C+,ZC3H13-,ZCCHC8+,ZFC3H1+,ZFP3-,ZMYM4-,ZNF141+,ZNF264+,ZNF280C-,ZNF292++,ZNF345+,ZNF383+,ZNF431+,ZNF441-,ZNF442-,ZNF484+,ZNF502-,ZNF546+,ZNF585A-,ZNF627--,ZNF639+,ZNF737-,ZRANB3- |
| **blue** | BP | GO:0006725 | cellular aromatic compound metabolic process | 1025 | 5017 | 3.30E-24 | 3.03E-21 | ADAL--,ADAT2-,AFG1L-,AHCTF1+,AIMP1-,ANKRD31-,ARMCX3-,ASF1A-,ATF6-,ATIC-,ATP5MG--,ATP5PB--,ATR-,ATRX+,BLM+,BLZF1-,BMI1+,BMT2+,BPNT1-,BRWD1+,CBX5+,CDK13+,CETN2-,CHD1+,CHURC1--,CLNS1A-,CLOCK+,CLPX-,CNOT10--,CPSF3-,CREBRF+,CREG1-,CRLF3+,CTH-,DCAF13-,DEK+,DNAJC2+,DUS4L-,ECD-,EGLN1+,EIF2A-,ELOC-,EPC1+,EPC2++,EPRS1-,ERH-,ETV1-,FANCF-,FASTKD3-,FIP1L1-,FOXO3+,G3BP1+,GALR1-,GART-,GEMIN2-,GNPNAT1+,GPD2-,GTF2A2-,GTF2B-,GTF2E1-,GTF2H3-,GTF2H5-,H4C3-,HIGD1A-,HMGN1-,HSD17B12-,IK-,INO80D+,INTS6+,JAK2+,LCOR++,LCORL+,LIG4-,LIN9+,LRRK2-,LSM3-,MAGOH-,MAP3K2+,MBD2+,ME1-,ME2-,MED4-,MEIS2-,METTL15-,MFAP1-,MIER3+,MLF1-,MMUT-,MPC1--,MRPS9--,MTR-,NAA16++,NBDY-,NCOA4-,NDUFA12-,NDUFC1-,NDUFS4-,NFAT5+,NFE2L3+,NIPSNAP2-,NUDT12-,NUDT4+,NUP42-,NUP54-,ORC3-,OTUD4+,PA2G4-,PDHB-,PDHX-,PHF3+,PLRG1-,POLI-,POLR2K--,POU5F2+,PPP4R2+,PRDX3-,PRKDC-,PRMT3-,PRMT6-,PRPF38B+,PRPF40A+,PRPF4B+,PSMD10-,PSMD14-,PURB+,PUS7+,QTRT2-,RAC1-,RBBP6+,RBMXL1-,RC3H2+,REL+,RESF1+,RHOQ+,RIOK1-,RNF14-,RNF168+,RORA+,RPF1-,SAP18-,SCARNA7+,SDHD-,SETD6+,SETDB2+,SF3B6-,SIRT1+,SLC35A3+,SLIRP-,SMAD4+,SMARCA1++,SMC5+,SNRPD1-,SNRPE-,SNW1-,SREK1+,SRSF10+,SUCLG1-,SUPT3H-,TATDN1-,TATDN3-,TDRD3-,TENT2-,TENT4B+,THOC2+,THOC7-,TIPIN-,TOPORS+,TRIP12-,TRPC1-,TSN-,TTC21B+,TUT4+,UFL1-,WDHD1--,XRCC4-,YY1+,ZBTB1+,ZBTB44+,ZC3H12C+,ZC3H13-,ZCCHC8+,ZFC3H1+,ZFP3-,ZMYM4-,ZNF141+,ZNF264+,ZNF280C-,ZNF292++,ZNF345+,ZNF383+,ZNF431+,ZNF441-,ZNF442-,ZNF484+,ZNF502-,ZNF546+,ZNF585A-,ZNF627--,ZNF639+,ZNF737-,ZRANB3- |
| **blue** | MF | GO:1901363 | heterocyclic compound binding | 1018 | 4946 | 8.17E-24 | 6.67E-21 | ABCD3-,AFG1L-,AGPS-,AHCTF1+,AIMP1-,ALDH1A1-,ATF6-,ATR-,ATRX+,BLM+,BLZF1-,CAPRIN1-,CDK13+,CDKL3-,CHD1+,CLNS1A-,CLOCK+,CLPX-,CPSF3-,CREBRF+,CRLF3+,CTH-,DBT-,DCAF13-,DDX50-,DEK+,DGKH-,DNAJC2+,DUS4L-,EIF2A- |

| | | | | | | | | |
|---|---|---|---|---|---|---|---|---|
| | | | | | | | | ,EIF2B1-,EIF2S1-,EIF2S3-,EIF3M-,EIF4E-,EPRS1-,ERH-,ETV1-,FASTKD3-,FIP1L1-,FOXO3+,G3BP1+,GART-,GDI2-,GFM1-,GNL2-,GTF2B-,GTF2H5-,GVINP1--,H4C3-,HMGN1-,JAK2+,LCOR++,LCORL+,LIG4-,LIN9+,LONP2-,LRRK2-,LSM3-,MAGOH-,MAK16-,MAP3K2+,MBD2+,ME1-,ME2-,MEIS2-,MFAP1-,MIER3+,MLF1-,MMUT-,MORC3+,MRPL13-,MRPS18C-,MRPS28-,MRPS35-,MRPS9--,MTR-,NFAT5+,NFE2L3+,NUDT4+,NUP42-,ORC3-,OTUD4+,PA2G4-,PDIA3-,PIN4-,PLPBP-,POLI-,POLR2K--,POU5F2+,PRKDC-,PRMT3-,PRPF38B+,PRPF40A+,PRPF4B+,PURB+,PUS7+,RABL3-,RAC1-,RBBP6+,RBM48+,RBMXL1-,RC3H2+,REL+,RHEB-,RHOQ+,RIOK1-,RORA+,RPF1-,SAP18-,SAR1A-,SCP2-,SDHD-,SETDB2+,SF3B6-,SIRT1+,SLFN5-,SLIRP-,SMAD4+,SMARCA1++,SMC5+,SNRPD1-,SNRPE-,SNW1-,SREK1+,SRP19+,SRP54-,SRP9-,SRSF10+,SUCLG1-,SUPT3H-,TBCA-,TDRD3-,TENT2-,TENT4B+,THOC2+,THOC7-,TIPIN-,TLK1-,TOPORS+,TSN-,TUT4+,UBE2E1-,WDHD1--,YWHAE-,YY1+,ZBTB1+,ZBTB44+,ZC3H13-,ZCCHC7-,ZCCHC8+,ZCCHC9-,ZFC3H1+,ZFP3-,ZMYM4-,ZNF141+,ZNF264+,ZNF280C-,ZNF292++,ZNF345+,ZNF383+,ZNF431+,ZNF441-,ZNF442-,ZNF484+,ZNF502-,ZNF546+,ZNF585A-,ZNF627--,ZNF639+,ZNF737-,ZRANB3- |
| **blue** | BP | GO:0010467 | gene expression | 935 | 4514 | 4.54E-23 | 3.33E-20 | ADAT2-,AHCTF1+,AIMP1-,ARMCX3-,ASF1A-,ATF6-,ATRX+,BIRC3+,BLM+,BLZF1-,BMI1+,BMT2+,BRWD1+,CAPRIN1-,CBX5+,CDK13+,CHD1+,CHURC1--,CIAO2A-,CLNS1A-,CLOCK+,CNOT10--,CNPY2--,CPSF3-,CREBRF+,CREG1-,CRLF3+,CTH-,DCAF13-,DEK+,DNAJB11-,DNAJC2+,DUS4L-,ECD-,EGLN1+,EIF2A-,EIF2B1-,EIF2S1-,EIF2S3-,EIF3M-,EIF4E-,ELOC-,EPC1+,EPC2++,EPRS1-,ETV1-,FASTKD3-,FIP1L1-,FOXO3+,GALR1-,GEMIN2-,GFM1-,GTF2A2-,GTF2B-,GTF2E1-,GTF2H3-,GTF2H5-,H4C3-,HIGD1A-,HMGN1-,IK-,IMMP1L-,INTS6+,ISCA2-,ITGB8+,JAK2+,LCOR++,LCORL+,LIN9+,LONP2-,LRRK2-,LSM3-,MAGOH-,MAP3K2+,MBD2+,MED4-,MEIS2-,METTL15-,MFAP1-,MIER3+,MLF1-,MRPL13-,MRPL47-,MRPL51-,MRPS18C-,MRPS28-,MRPS35-,MRPS9--,NAA16++,NBDY-,NCOA4-,NF1-,NFAT5+,NFE2L3+,NUP42-,NUP54-,PA2G4-,PHF3+,PLGRKT-,PLRG1-,POLR2K--,POU5F2+,PPP4R2+,PRDX3-,PRKDC-,PRMT3-,PRMT6-,PRPF38B+,PRPF40A+,PRPF4B+,PSMD10-,PSMD14-,PURB+,PUS7+,QTRT2-,RBBP6+,RBMXL1-,RC3H2+,REL+,RESF1+,RHOQ+,RIOK1-,RNF14-,RNF168+,RORA+,RPF1-,SAP18-,SCARNA7+,SETD6+,SETDB2+,SF3B6-,SIRT1+,SMAD4+,SMARCA1++,SNRPD1-,SNRPE-,SNW1-,SPCS2-,SREK1+,SRP9-,SRSF10+,SUPT3H-,TENT2-,TENT4B+,THOC2+,THOC7-,TMA7-,TOPORS+,TSN-,TTC21B+,TUT4+,UFL1-,WDHD1--,YY1+,ZBTB1+,ZBTB44+,ZC3H13-,ZCCHC8+,ZFC3H1+,ZFP3-,ZMYM4-,ZNF141+,ZNF264+,ZNF280C-,ZNF292++,ZNF345+,ZNF383+,ZNF431+,ZNF441-,ZNF442-,ZNF484+,ZNF502-,ZNF546+,ZNF585A-,ZNF627--,ZNF639+,ZNF737- |
| **cyan** | BP | GO:0006119 | oxidative phosphorylation | 26 | 122 | 1.96E-13 | 1.04E-09 | ATP5F1A-,ATP5F1B--,ATP5F1C--,ATP5F1E-,ATP5MC3-,ATP5PF-,ATP5PO-,COX15--,COX5A-,COX7A2-,COX7B-,COX7C-,NDUFA1--,NDUFA4-,NDUFA6--,NDUFAB1-,NDUFAF1-,NDUFB1--,NDUFB3--,NDUFB4-,PARK7-,UQCRC2-- |
| **cyan** | BP | GO:0009144 | purine nucleoside triphosphate metabolic process | 33 | 291 | 4.17E-13 | 1.04E-09 | ATP5F1A-,ATP5F1B--,ATP5F1C--,ATP5F1E-,ATP5MC3-,ATP5PF-,ATP5PO-,BPGM--,COX15--,COX5A-,COX7A2-,COX7B-,COX7C-,DGUOK-,NDUFA1--,NDUFA4-,NDUFA6--,NDUFAB1-,NDUFAF1-,NDUFB1--,NDUFB3--,NDUFB4-,PARK7-,PGAM1-,PRKAG1--,UQCRC2-- |
| **cyan** | BP | GO:0046034 | ATP metabolic process | 31 | 257 | 4.24E-13 | 1.04E-09 | ATP5F1A-,ATP5F1B--,ATP5F1C--,ATP5F1E-,ATP5MC3-,ATP5PF-,ATP5PO-,BPGM--,COX15--,COX5A-,COX7A2-,COX7B-,COX7C-,NDUFA1--,NDUFA4-,NDUFA6--,NDUFAB1-,NDUFAF1-,NDUFB1--,NDUFB3--,NDUFB4-,PARK7-,PGAM1-,PRKAG1--,UQCRC2-- |
| **cyan** | BP | GO:0044281 | small molecule metabolic process | 76 | 1709 | 6.47E-13 | 1.19E-09 | ATP5F1A-,ATP5F1B--,ATP5F1C--,ATP5F1E-,ATP5MC3-,ATP5PF-,ATP5PO-,BPGM--,CHP1-,COX15--,COX5A-,COX7A2-,COX7B-,COX7C-,DGUOK-,FARSB-,FH-,FUCA1-,GSTO1-,HEXB-,KARS1-,LCMT1-,LGMN-,MGST3-,NDUFA1--,NDUFA4-,NDUFA6--,NDUFAB1-,NDUFAF1-,NDUFB1--,NDUFB3--,NDUFB4-,OSBP-,PARK7-,PECR-,PGAM1-,PPCS-,PPT1--,PRKAG1--,PSMB2-,PSMB4-,PSMC2-,PSMD13-,PUDP-,RDH11-,RRM1-,SARS1-,SDHB-,SRR--,UQCRC2--,VDAC1- |

| | | | | | | | | |
|---|---|---|---|---|---|---|---|---|
| **cyan** | BP | GO:0009205 | purine ribonucleoside triphosphate metabolic process | 32 | 285 | 1.32E-12 | 1.68E-09 | ATP5F1A-,ATP5F1B--,ATP5F1C--,ATP5F1E-,ATP5MC3-,ATP5PF-,ATP5PO-,BPGM--,COX15--,COX5A-,COX7A2-,COX7B-,COX7C-,NDUFA1--,NDUFA4-,NDUFA6--,NDUFAB1-,NDUFAF1-,NDUFB1--,NDUFB3--,NDUFB4-,PARK7-,PGAM1-,PRKAG1--,UQCRC2-- |
| **cyan** | BP | GO:0009123 | nucleoside monophosphate metabolic process | 33 | 321 | 1.57E-12 | 1.68E-09 | ATP5F1A-,ATP5F1B--,ATP5F1C--,ATP5F1E-,ATP5MC3-,ATP5PF-,ATP5PO-,BPGM--,COX15--,COX5A-,COX7A2-,COX7B-,COX7C-,DGUOK-,NDUFA1--,NDUFA4-,NDUFA6--,NDUFAB1-,NDUFAF1-,NDUFB1--,NDUFB3--,NDUFB4-,PARK7-,PGAM1-,PRKAG1--,UQCRC2-- |
| **cyan** | BP | GO:0009141 | nucleoside triphosphate metabolic process | 33 | 309 | 1.91E-12 | 1.68E-09 | ATP5F1A-,ATP5F1B--,ATP5F1C--,ATP5F1E-,ATP5MC3-,ATP5PF-,ATP5PO-,BPGM--,COX15--,COX5A-,COX7A2-,COX7B-,COX7C-,DGUOK-,NDUFA1--,NDUFA4-,NDUFA6--,NDUFAB1-,NDUFAF1-,NDUFB1--,NDUFB3--,NDUFB4-,PARK7-,PGAM1-,PRKAG1--,UQCRC2-- |
| **cyan** | BP | GO:0009199 | ribonucleoside triphosphate metabolic process | 32 | 291 | 2.01E-12 | 1.68E-09 | ATP5F1A-,ATP5F1B--,ATP5F1C--,ATP5F1E-,ATP5MC3-,ATP5PF-,ATP5PO-,BPGM--,COX15--,COX5A-,COX7A2-,COX7B-,COX7C-,NDUFA1--,NDUFA4-,NDUFA6--,NDUFAB1-,NDUFAF1-,NDUFB1--,NDUFB3--,NDUFB4-,PARK7-,PGAM1-,PRKAG1--,UQCRC2-- |
| **cyan** | BP | GO:0009161 | ribonucleoside monophosphate metabolic process | 32 | 304 | 2.06E-12 | 1.68E-09 | ATP5F1A-,ATP5F1B--,ATP5F1C--,ATP5F1E-,ATP5MC3-,ATP5PF-,ATP5PO-,BPGM--,COX15--,COX5A-,COX7A2-,COX7B-,COX7C-,NDUFA1--,NDUFA4-,NDUFA6--,NDUFAB1-,NDUFAF1-,NDUFB1--,NDUFB3--,NDUFB4-,PARK7-,PGAM1-,PRKAG1--,UQCRC2-- |
| **cyan** | BP | GO:0009167 | purine ribonucleoside monophosphate metabolic process | 31 | 291 | 4.47E-12 | 3.28E-09 | ATP5F1A-,ATP5F1B--,ATP5F1C--,ATP5F1E-,ATP5MC3-,ATP5PF-,ATP5PO-,BPGM--,COX15--,COX5A-,COX7A2-,COX7B-,COX7C-,NDUFA1--,NDUFA4-,NDUFA6--,NDUFAB1-,NDUFAF1-,NDUFB1--,NDUFB3--,NDUFB4-,PARK7-,PGAM1-,PRKAG1--,UQCRC2-- |
| **Dark grey** | MF | GO:0140110 | transcription regulator activity | 8 | 1623 | 7.05E-05 | 0.31 | TCP10L+,ZNF749+ |
| **Dark grey** | MF | GO:0000981 | DNA-binding transcription factor activity, RNA polymerase II-specific | 7 | 1209 | 8.51E-05 | 0.31 | ZNF749+ |
| **Dark grey** | MF | GO:0003700 | DNA-binding transcription factor activity | 7 | 1298 | 0.00014 | 0.33 | ZNF749+ |
| **Dark grey** | BP | GO:0006357 | regulation of transcription by RNA polymerase II | 8 | 2123 | 0.00049 | 0.90 | TCP10L+,ZNF749+ |
| **Dark grey** | BP | GO:0006366 | transcription by RNA polymerase II | 8 | 2250 | 0.00075 | 1 | TCP10L+,ZNF749+ |
| **Dark turquoise** | MF | GO:0043138 | 3'-5' DNA helicase activity | 2 | 19 | 0.0025 | 1 | |
| **Dark turquoise** | MF | GO:0016787 | hydrolase activity | 17 | 1995 | 0.0035 | 1 | METAP1D-,RAB4B-,RGS11-,USP31+ |
| **Dark turquoise** | MF | GO:0003824 | catalytic activity | 31 | 4682 | 0.0038 | 1 | METAP1D-,PDIA2-,RAB4B-,RGS11-,RNF130+,USP31+ |
| **Dark turquoise** | MF | GO:0004843 | thiol-dependent ubiquitin-specific protease activity | 3 | 84 | 0.0040 | 1 | USP31+ |
| **Dark turquoise** | MF | GO:0036459 | thiol-dependent ubiquitinyl hydrolase activity | 3 | 92 | 0.0054 | 1 | USP31+ |
| **Green yellow** | BP | GO:0050896 | response to stimulus | 293 | 6519 | 7.93E-17 | 5.37E-13 | ABCG2-,AKR1C1+,ANO6+,BMP6-,C7+,CASP7+,CASQ2-,CCND1-,CD58+,CFI+,CLEC2B+,CLEC5A++,CTNND1-,EPS8-,FBXO32+,FOXN3+,GBP2+,HEBP2++,HIPK1+,IFI16+,IFNAR2-,IL13RA1+,IQGAP2+,KDR-,LEPROT-,LYZ+,MAML2+,MYL9-,NEDD4++,NPNT+,PRRX1+,PYGL+,RAB13+,RAB29+,RASSF3++,RECQL- |

| | | | | | | | | |
|---|---|---|---|---|---|---|---|---|
| | | | | | | | | ,S100A4+,SLC40A1-,SLC7A2++,SMOC2-,STK38+,SWAP70+,TRIB2-,UBR5+,WWTR1++,YAP1+ |
| **Green yellow** | BP | GO:0030198 | extracellular matrix organization | 43 | 287 | 1.46E-16 | 5.37E-13 | COL13A1-,COL14A1-,KDR-,LAMC1+,NID2-,NPNT+,SMOC2- |
| **Green yellow** | BP | GO:0002376 | immune system process | 133 | 2121 | 2.59E-16 | 6.35E-13 | ANO6+,BMP6-,C7+,CD58+,CFI+,CLEC2B+,CLEC5A++,EPS8-,GBP2+,HEBP2++,HIPK1+,IFI16+,IFNAR2-,IQGAP2+,KDR-,LYZ+,MYL9-,PYGL+,RAB29+,SLC40A1-,SLC7A2++,SWAP70+,YAP1+ |
| **Green yellow** | MF | GO:0005201 | extracellular matrix structural constituent | 29 | 128 | 6.46E-16 | 1.19E-12 | COL13A1-,COL14A1-,LAMC1+,NID2-,NPNT+ |
| **Green yellow** | BP | GO:0043062 | extracellular structure organization | 44 | 321 | 1.44E-15 | 2.12E-12 | COL13A1-,COL14A1-,KDR-,LAMC1+,NID2-,NPNT+,SMOC2- |
| **Green yellow** | BP | GO:0006955 | immune response | 95 | 1399 | 7.84E-14 | 9.60E-11 | ANO6+,BMP6-,C7+,CD58+,CFI+,CLEC2B+,CLEC5A++,GBP2+,HEBP2++,IFI16+,IFNAR2-,IQGAP2+,LYZ+,PYGL+,RAB29+,SWAP70+ |
| **Green yellow** | BP | GO:0007166 | cell surface receptor signaling pathway | 133 | 2245 | 1.98E-13 | 2.08E-10 | ANO6+,BMP6-,CCND1-,CTNND1-,GBP2+,HIPK1+,IFNAR2-,IL13RA1+,KDR-,LEPROT-,MAML2+,NEDD4++,NPNT+,PRRX1+,RAB29+,SMOC2-,UBR5+,WWTR1++,YAP1+ |
| **Green yellow** | BP | GO:0007155 | cell adhesion | 83 | 1111 | 9.87E-13 | 9.06E-10 | BMP6-,CD58+,COL13A1-,COL14A1-,CTNND1-,KDR-,LAMC1+,LPP+,MYL9-,NID2-,NPNT+,PCDH18+,SMOC2-,SWAP70+ |
| **Green yellow** | BP | GO:0022610 | biological adhesion | 83 | 1115 | 1.16E-12 | 9.44E-10 | BMP6-,CD58+,COL13A1-,COL14A1-,CTNND1-,KDR-,LAMC1+,LPP+,MYL9-,NID2-,NPNT+,PCDH18+,SMOC2-,SWAP70+ |
| **Green yellow** | BP | GO:0070887 | cellular response to chemical stimulus | 139 | 2502 | 1.68E-12 | 1.24E-09 | ABCG2-,AKR1C1+,ANO6+,BMP6-,CASP7+,CASQ2-,CCND1-,CD58+,EPS8-,FBXO32+,GBP2+,HIPK1+,IFI16+,IFNAR2-,IL13RA1+,KDR-,LEPROT-,NEDD4++,NPNT+,RAB13+,SLC40A1-,SMOC2-,SWAP70+,UBR5+,YAP1+ |
| **Light yellow** | BP | GO:0051094 | positive regulation of developmental process | 21 | 1088 | 3.48E-05 | 0.072 | BMPR1A+,BTG1++,CCDC71L+,FBXO5+,HGF+,KL-,MALT1+,SOCS2+ |
| **Light yellow** | BP | GO:0019220 | regulation of phosphate metabolic process | 24 | 1403 | 5.33E-05 | 0.072 | BMPR1A+,DUSP6-,HGF+,KL-,MALT1+,PDK1+,SOCS2+ |
| **Light yellow** | BP | GO:0051174 | regulation of phosphorus metabolic process | 24 | 1405 | 5.45E-05 | 0.072 | BMPR1A+,DUSP6-,HGF+,KL-,MALT1+,PDK1+,SOCS2+ |
| **Light yellow** | BP | GO:0045597 | positive regulation of cell differentiation | 17 | 785 | 5.45E-05 | 0.072 | BMPR1A+,BTG1++,CCDC71L+,FBXO5+,HGF+,MALT1+,SOCS2+ |
| **Light yellow** | BP | GO:0042327 | positive regulation of phosphorylation | 17 | 790 | 5.83E-05 | 0.072 | BMPR1A+,DUSP6-,HGF+,KL-,MALT1+ |
| **magenta** | MF | GO:0000981 | DNA-binding transcription factor activity, RNA polymerase II-specific | 53 | 1209 | 1.96E-08 | 0.00011 | FOXD4L5+,MLLT3+,RFX7+,ZNF285-,ZNF550+,ZNF776+,ZNF829++,ZSCAN23- |
| **magenta** | MF | GO:0003700 | DNA-binding transcription factor activity | 55 | 1298 | 2.97E-08 | 0.00011 | FOXD4L5+,GOLGB1+,MLLT10+,MLLT3+,RFX7+,ZNF285-,ZNF550+,ZNF776+,ZNF829++,ZSCAN23- |
| **magenta** | MF | GO:0003677 | DNA binding | 68 | 1976 | 1.59E-06 | 0.0029 | CENPC+,CXXC4+,FOXD4L5+,GOLGB1+,MLLT10+,REV3L+,RFX7+,SHPRH+,ZNF285-,ZNF550+,ZNF776+,ZNF829++,ZSCAN23- |
| **magenta** | MF | GO:0140110 | transcription regulator activity | 59 | 1623 | 1.92E-06 | 0.0029 | FOXD4L5+,GOLGB1+,MLLT10+,MLLT3+,RFX7+,ZNF285-,ZNF550+,ZNF776+,ZNF829++,ZSCAN23- |
| **magenta** | BP | GO:0044782 | cilium organization | 21 | 324 | 1.98E-06 | 0.0029 | CEP126+,CFAP44+,HAUS3+ |
| **magenta** | BP | GO:0060271 | cilium assembly | 20 | 309 | 3.51E-06 | 0.0043 | CEP126+,CFAP44+,HAUS3+ |
| **magenta** | BP | GO:0061512 | protein localization to cilium | 7 | 47 | 3.48E-05 | 0.036 | |
| **pink** | BP | GO:0048709 | oligodendrocyte differentiation | 17 | 88 | 1.92E-08 | 0.00014 | DAAM2-,HDAC11-,NKX6-2+ |
| **pink** | BP | GO:0010001 | glial cell differentiation | 24 | 189 | 1.29E-07 | 0.00047 | DAAM2-,HDAC11-,NKX6-2+,SIRT2- |
| **pink** | BP | GO:0042063 | gliogenesis | 27 | 246 | 4.66E-07 | 0.0011 | DAAM2-,E2F1-,HDAC11-,NKX6-2+,SIRT2-,SUN2+ |
| **pink** | BP | GO:0014013 | regulation of gliogenesis | 15 | 97 | 2.66E-06 | 0.0049 | DAAM2-,E2F1-,NKX6-2+,SIRT2- |

| | | | | | | | | |
|---|---|---|---|---|---|---|---|---|
| **pink** | BP | GO:0042552 | myelination | 16 | 121 | 1.22E-05 | 0.016 | MAL-,NKX6-2+,PLLP-,SIRT2- |
| **pink** | BP | GO:0007272 | ensheathment of neurons | 16 | 123 | 1.52E-05 | 0.016 | MAL-,NKX6-2+,PLLP-,SIRT2- |
| **pink** | BP | GO:0008366 | axon ensheathment | 16 | 123 | 1.52E-05 | 0.016 | MAL-,NKX6-2+,PLLP-,SIRT2- |
| **pink** | BP | GO:0014003 | oligodendrocyte development | 9 | 42 | 1.70E-05 | 0.016 | HDAC11-,NKX6-2+ |
| **pink** | BP | GO:0014015 | positive regulation of gliogenesis | 10 | 60 | 5.50E-05 | 0.039 | E2F1-,NKX6-2+ |
| **pink** | BP | GO:0045685 | regulation of glial cell differentiation | 10 | 60 | 6.24E-05 | 0.039 | DAAM2-,NKX6-2+ |
| **salmon** | BP | GO:0043933 | protein-containing complex subunit organization | 74 | 1926 | 1.47E-10 | 1.08E-06 | ABT1-,AIMP2-,BBS4-,CCDC115-,CHMP4B-,COA3--,COX14-,DDB1-,EIF3C-,EIF3K-,EIF3L-,EIF4H-,GBA-,GLE1-,H2BC15-,HAX1--,MRPL11-,MRPL2-,MRPL27--,MRPL37-,MRPL54-,MRPL57-,MSRB2-,NDUFA2--,NDUFA8-,NDUFB2--,NDUFB8--,NDUFS3--,NDUFS6-,NFS1--,NME2-,PDCD6-,PDZD11--,PFDN6-,PSMC3--,PSMD4-,SAMM50-,STOML2-,TRAPPC1--,TRAPPC4-,UQCC1-,UQCR10-,VIPAS39- |
| **salmon** | BP | GO:0034622 | cellular protein-containing complex assembly | 49 | 985 | 3.44E-10 | 1.26E-06 | ABT1-,BBS4-,CCDC115-,COA3--,COX14-,DDB1-,EIF3C-,EIF3K-,EIF3L-,EIF4H-,H2BC15-,HAX1--,MRPL11-,MSRB2-,NDUFA2--,NDUFA8-,NDUFB2--,NDUFB8--,NDUFS3--,NDUFS6-,PFDN6-,PSMC3--,PSMD4-,SAMM50-,UQCC1-,UQCR10- |
| **salmon** | BP | GO:0065003 | protein-containing complex assembly | 64 | 1645 | 6.09E-10 | 1.49E-06 | ABT1-,AIMP2-,BBS4-,CCDC115-,CHMP4B-,COA3--,COX14-,DDB1-,EIF3C-,EIF3K-,EIF3L-,EIF4H-,GBA-,H2BC15-,HAX1--,MRPL11-,MSRB2-,NDUFA2--,NDUFA8-,NDUFB2--,NDUFB8--,NDUFS3--,NDUFS6-,NFS1--,NME2-,PDCD6-,PDZD11--,PFDN6-,PSMC3--,PSMD4-,SAMM50-,STOML2-,TRAPPC1--,TRAPPC4-,UQCC1-,UQCR10- |
| **salmon** | BP | GO:0016043 | cellular component organization | 130 | 5383 | 4.51E-09 | 8.27E-06 | ABT1-,ACTG1-,AIMP2-,ALAS1-,ANAPC15--,ATP5MC1-,ATP5MF--,BABAM2-,BBS4-,CAPNS1-,CCDC115-,CHMP4B-,COA3--,COPS6-,COX14-,DDB1-,EIF3C-,EIF3K-,EIF3L-,EIF4H-,FUNDC2-,GBA-,GLE1-,H2BC15-,HAX1--,IFT27--,IQGAP3--,MAP2K5--,MRPL11-,MRPL2-,MRPL27--,MRPL37-,MRPL54-,MRPL57-,MSRB2-,NDUFA2--,NDUFA8-,NDUFB2--,NDUFB8--,NDUFS3--,NDUFS6-,NFS1--,NHP2-,NME2-,NPM3--,PDCD6-,PDZD11--,PEX19-,PEX7-,PFDN6-,PHB2-,PSMC3--,PSMD4-,RAB5B-,SAMM50-,SLC25A5--,STOML2-,STRADA-,SUPT4H1-,TCTN3--,TRAPPC1--,TRAPPC4-,TTC21A-,UQCC1-,UQCR10-,VIPAS39-,VPS25--,VPS33B-,VPS39- |
| **salmon** | BP | GO:0071840 | cellular component organization or biogenesis | 134 | 5565 | 6.72E-09 | 9.86E-06 | ABT1-,ACTG1-,AIMP2-,ALAS1-,ANAPC15--,ATP5MC1-,ATP5MF--,BABAM2-,BBS4-,CAPNS1-,CCDC115-,CHMP4B-,COA3--,COPS6-,COX14-,DDB1-,EIF3C-,EIF3K-,EIF3L-,EIF4H-,FUNDC2-,GBA-,GLE1-,H2BC15-,HAX1--,IFT27--,IQGAP3--,MAP2K5--,MRPL11-,MRPL2-,MRPL27--,MRPL37-,MRPL54-,MRPL57-,MSRB2-,NDUFA2--,NDUFA8-,NDUFB2--,NDUFB8--,NDUFS3--,NDUFS6-,NFS1--,NHP2-,NME2-,NPM3--,PDCD6-,PDZD11--,PEX19-,PEX7-,PFDN6-,PHB2-,POP4-,PSMC3--,PSMD4-,RAB5B-,SAMM50-,SLC25A5--,STOML2-,STRADA-,SUPT4H1-,TCTN3--,TRAPPC1--,TRAPPC4-,TTC21A-,UQCC1-,UQCR10-,VIPAS39-,VPS25--,VPS33B-,VPS39- |
| **salmon** | BP | GO:0006119 | oxidative phosphorylation | 19 | 122 | 2.10E-08 | 2.57E-05 | ATP5MC1-,ATP5MF--,COX4I1--,COX5B-,COX6B1--,NDUFA2--,NDUFA8-,NDUFB2--,NDUFB8--,NDUFS3--,NDUFS6-,STOML2-,UQCR10-,UQCRQ- |
| **salmon** | BP | GO:0044085 | cellular component biogenesis | 83 | 2846 | 5.61E-08 | 5.56E-05 | ABT1-,ACTG1-,AIMP2-,BBS4-,CCDC115-,CHMP4B-,COA3--,COX14-,DDB1-,EIF3C-,EIF3K-,EIF3L-,EIF4H-,GBA-,H2BC15-,HAX1--,IFT27--,MRPL11-,MSRB2-,NDUFA2--,NDUFA8-,NDUFB2--,NDUFB8--,NDUFS3--,NDUFS6-,NFS1--,NHP2-,NME2-,NPM3--,PDCD6-,PDZD11--,PFDN6-,POP4-,PSMC3--,PSMD4-,SAMM50-,STOML2-,TCTN3--,TRAPPC1--,TRAPPC4-,UQCC1-,UQCR10-,VPS25-- |
| **salmon** | BP | GO:0022607 | cellular component assembly | 76 | 2610 | 6.05E-08 | 5.56E-05 | ABT1-,ACTG1-,AIMP2-,BBS4-,CCDC115-,CHMP4B-,COA3--,COX14-,DDB1-,EIF3C-,EIF3K-,EIF3L-,EIF4H-,GBA-,H2BC15-,HAX1--,IFT27--,MRPL11-,MSRB2-,NDUFA2--,NDUFA8-,NDUFB2--,NDUFB8--,NDUFS3--,NDUFS6-,NFS1--,NME2-,PDCD6-,PDZD11--,PFDN6-,PSMC3--,PSMD4-,SAMM50-,STOML2-,TCTN3--,TRAPPC1--,TRAPPC4-,UQCC1-,UQCR10-,VPS25-- |
| **salmon** | BP | GO:1901564 | organonitrogen compound metabolic process | 135 | 5499 | 4.17E-07 | 0.00034 | AARSD1-,AHCY-,AIMP2-,AKR1A1-,ALAS1-,ALDH18A1-,ANAPC15--,ATP5MC1-,ATP5MF--,BABAM2-,BPHL-,CAPNS1-,CCDC115-,CHMP4B-,COA3--,COMMD1-,COPS6-,COX4I1--,COX5B-,COX6B1--,DDB1-,DDOST-,EEF1AKNMT-,EIF3C-,EIF3K-,EIF3L-,EIF4H- |

| | | | | | | | | |
|---|---|---|---|---|---|---|---|---|
| | | | | | | | | ,ERP29-,ESYT1-,GBA-,GLE1-,GMPR2-,GSS-,H2BC15-,HAX1--,HMOX2-,IQGAP3--,MAP2K5--,MED8-,MRPL11-,MRPL2-,MRPL27--,MRPL37-,MRPL54-,MRPL57-,MSRB2-,NDUFA2--,NDUFA8-,NDUFB2--,NDUFB8--,NDUFS3--,NDUFS6-,NFS1--,NHLRC1-,NME2-,PDCD6-,PDZD11--,PEBP1--,PHB2-,PSMC3--,PSMD4-,PSMD8-,PSME3--,STOML2-,STRADA-,UQCC1-,UQCR10-,UQCRQ-,VIPAS39-,VPS25--,VPS33B-,ZC3HC1- |
| **salmon** | BP | GO:0006996 | organelle organization | 86 | 3315 | 6.22E-07 | 0.00046 | ABT1-,ACTG1-,ALAS1-,ANAPC15--,ATP5MC1-,ATP5MF--,BABAM2-,BBS4-,CCDC115-,CHMP4B-,COA3--,COPS6-,COX14-,DDB1-,FUNDC2-,GBA-,H2BC15-,HAX1--,IFT27--,MRPL11-,MSRB2-,NDUFA2--,NDUFA8-,NDUFB2--,NDUFB8--,NDUFS3--,NDUFS6-,NHP2-,NPM3--,PDCD6-,PEX19-,PEX7-,PHB2-,RAB5B-,SAMM50-,SLC25A5--,STOML2-,SUPT4H1-,TCTN3--,TRAPPC1--,TRAPPC4-,TTC21A-,UQCC1-,UQCR10-,VPS25--,VPS33B-,VPS39- |
| **turquoise** | BP | GO:0044238 | primary metabolic process | 1543 | 8441 | 3.21E-06 | 0.012 | AAAS-,AARS2-,ABHD14B-,ACADS-,ACAN+,ACOT8+,ACSS1-,ADAM15-,ADAM8-,ADAMTS13-,ADAMTS2++,ADCYAP1R1+,ADIPOR1--,ADPGK-,AGPAT1--,AKR7A2-,ALDH4A1-,ALYREF-,ANKRD9+,APH1A-,APLNR+,APOE-,ARHGEF10L+,ASPHD1-,ATAT1-,ATF5-,ATF6B-,ATG4B+,ATN1+,ATP5MC2-,AUTS2-,B3GALT4-,BCAP31-,BCKDHA-,BOLA1-,BRD1+,BRD2+,BRD4+,BRPF3+,CACNA1H-,CALR--,CASP9-,CBS+,CBY1-,CCDC88C-,CCS--,CES2-,CHST6++,CIB1-,CIC+,CINP-,CLEC3B-,CNDP2-,COMT-,COQ9-,CRAT-,CREBBP+,CRYL1-,CSNK2B-,CST3-,CSTB-,CYB5R3-,DCAF11-,DCAKD-,DDX49-,DELE1-,DESI1-,DGCR8+,DHRS7B-,DHX8-,DIDO1+,DLST-,DOK7+,DUS2-,ECH1-,EDEM2--,EEF2-,EEF2K+,EIF3D-,EIF5A-,ELOB-,ETFB--,FAH-,FAM3A-,FANCE+,FBXL12-,FBXO17+,FBXO6-,FBXW8-,FGFR3-,FIS1-,FKBP2-,FKBP9-,FLAD1-,FLCN+,FMC1-,FN3KRP-,FSTL3+,G6PC3-,G6PD-,GAMT-,GANAB-,GCDH-,GLI1+,GLMP-,GORASP1-,GPAT4+,GPR37L1-,GPX4-,GSTP1-,GTF2H4-,GUK1-,GYS1-,H2BC4-,HDAC4++,HES4--,HES5-,HINT2--,HMBS-,HMGCL-,HNRNPL-,HR-,HRAS-,IKZF4+,IMP4-,INPP5K-,INTS3--,IRAK2++,IRF2BPL+,IRS2++,ITIH4-,IVD-,JADE2+,JARID2+,JDP2+,JUND+,KCTD5-,KDM4B+,KDM6B+,KLHL21+,LAMTOR1-,LARS2--,LSM7-,LTBP4+,MAPT-AS1+,MCM3-,MECP2+,MED20-,MFHAS1+,MGMT-,MICAL1-,MMP28--,MPV17L2-,MRM1-,MRPL20-,MRPL24--,MRPL53-,MRPS18B-,MSRB1--,MT3-,MTG2-,MYH3+,NCSTN-,NENF-,NFKBID+,NMRAL1-,NOL3-,NR6A1+,NTHL1-,NUCB1-,NUDT2--,NUP210-,OS9-,PARP3-,PARS2-,PC-,PDCD11-,PDE9A-,PER1+,PEX10-,PIM3+,PLK5+,PLPP4+,PMM2-,PNPLA6+,POLR2G-,POLR2L-,PPIF-,PPP1CA-,PPP1R1B-,PPP4C-,PRELID1-,PRKCD--,PSENEN--,PSMB10-,PTPN7-,PTPRU+,PUF60-,RAD9A-,RASA4+,RBM10-,RECQL5-,RERE+,RETSAT-,RFXANK-,RGMA+,RNF167-,RNF181-,RNF5-,RPN2-,RPS15-,RPS19-,RUBCN+,RUVBL1-,S100A1--,SARDH-,SCARNA13+,SDSL--,SEC13--,SF3A1--,SF3B5-,SH2D3C-,SHISA5-,SKI+,SLC35C2-,SLC44A2--,SMPD1-,SNAPIN-,SNHG12+,SNORA73A+,SNRNP40-,SOX12+,SPRED3-,SRRM2+,STARD10+,SUMF1-,SUMF2-,SURF1-,SWI5-,TAB1-,TBC1D10A-,TFPT-,TIMELESS+,TKFC-,TM7SF2--,TMEM258-,TMPRSS5+,TNFSF13-,TPP1-,TRAPPC9-,TRIM47+,TRIM8+,TRPM4-,TRRAP+,TSHZ1+,TSR2-,TST-,TXN2--,UBE2I+,UQCR11-,URB1+,USP4-,VKORC1-,VRK3-,WNT4+,XYLT2-,ZBED6++,ZBTB7C+,ZDHHC11+,ZDHHC4--,ZMIZ1+,ZNF16-,ZNF219+,ZNF358+,ZNF445+,ZNF496-,ZNF500+,ZNF511+,ZNF589+,ZNF750+,ZNF768+,ZNF853+,ZNHIT1-,ZSCAN25- |
| **turquoise** | BP | GO:0018205 | peptidyl-lysine modification | 93 | 354 | 5.72E-06 | 0.012 | AAAS-,ATAT1-,AUTS2-,BRD1+,BRD4+,BRPF3+,CREBBP+,FLCN+,HDAC4++,HINT2--,JADE2+,JARID2+,MECP2+,NUP210-,PER1+,RUVBL1-,TRPM4-,TRRAP+,UBE2I+,ZMIZ1+ |
| **turquoise** | BP | GO:0008152 | metabolic process | 1648 | 9084 | 6.43E-06 | 0.012 | AAAS-,AARS2-,ABHD14B-,ACADS-,ACAN+,ACOT8+,ACSS1-,ADAM15-,ADAM8-,ADAMTS13-,ADAMTS2++,ADCYAP1R1+,ADIPOR1--,ADPGK-,AGPAT1--,AKR7A2-,ALDH4A1-,ALKBH6-,ALYREF-,ANKRD9+,APH1A-,APLNR+,APOE-,ARAP1+,ARHGEF10L+,ASPHD1-,ATAT1-,ATF5-,ATF6B-,ATG4B+,ATN1+,ATP5MC2-,AUTS2-,B3GALT4-,BCAP31-,BCKDHA-,BOLA1-,BRD1+,BRD2+,BRD4+,BRPF3+,BSG-,CACNA1H-,CALR--,CASP9-,CBS+,CBY1-,CCDC88C-,CCS--,CES2-,CHMP2A-,CHST6++,CIB1-,CIC+,CINP-,CLEC3B-,CNDP2-,COMT-,COQ9-,CRAT-,CREBBP+,CRYL1-,CSNK2B-,CST3-,CSTB-,CYB5R3-,DCAF11-,DCAKD-,DDX49-,DELE1-,DESI1-,DGCR8+,DHRS7B-,DHX8-,DIDO1+,DLST-,DOK7+,DUS2-,ECH1-,EDEM2--,EEF2-,EEF2K+,EIF3D-,EIF5A-,ELOB-,ETFB--,FAH-,FAM3A-,FANCE+,FBXL12-,FBXO17+,FBXO6- |

| | | | | | | | | |
|---|---|---|---|---|---|---|---|---|
| | | | | | | | | ,FBXW8-,FGFR3-,FIS1-,FKBP2-,FKBP9-,FLAD1-,FLCN+,FMC1-,FN3KRP-,FSTL3+,G6PC3-,G6PD-,GABARAP-,GAMT-,GANAB-,GCDH-,GLI1+,GLMP-,GORASP1-,GPAT4+,GPR37L1-,GPX4-,GSTK1-,GSTP1-,GTF2H4-,GUK1-,GYS1-,H2BC4-,HDAC4++,HES4--,HES5-,HIGD2A-,HINT2--,HMBS-,HMGCL-,HNRNPL-,HR-,HRAS-,IKZF4+,IMP4-,INPP5K-,INTS3--,IRAK2++,IRF2BPL+,IRS2++,ITIH4-,IVD-,JADE2+,JARID2+,JDP2+,JUND+,KCTD5-,KDM4B+,KDM6B+,KLHL21+,LAMTOR1-,LAMTOR4-,LARS2--,LDHD-,LSM7-,LTBP4+,MAPT-AS1+,MCM3-,MECP2+,MED20-,MFHAS1+,MFN2-,MGMT-,MICAL1-,MICAL3+,MMP28--,MPV17-,MPV17L2-,MRM1-,MRPL20-,MRPL24--,MRPL53-,MRPS18B-,MSRB1--,MT3-,MTG2-,MYH3+,NCSTN-,NENF-,NFKBID+,NIT1-,NMRAL1-,NOL3-,NR6A1+,NTHL1-,NUCB1-,NUDT2--,NUP210-,OGFOD2-,OS9-,PARP3-,PARS2-,PC-,PDCD11-,PDE9A-,PER1+,PEX10-,PIM3+,PLK5+,PLPP4+,PMM2-,PNPLA6+,POLR2G-,POLR2L-,PPIF-,PPP1CA-,PPP1R1B-,PPP4C-,PRELID1-,PRKCD--,PSENEN--,PSMB10-,PTPN7-,PTPRU+,PUF60-,RAD9A-,RASA4+,RBM10-,RECQL5-,RERE+,RETSAT-,RFXANK-,RGMA+,RNF167-,RNF181-,RNF5-,ROMO1-,RPN2-,RPS15-,RPS19-,RUBCN+,RUVBL1-,S100A1--,SARDH-,SCARNA13+,SDSL--,SEC13--,SF3A1--,SF3B5-,SH2D3C-,SH3BGRL3-,SHISA5-,SKI+,SLC35C2-,SLC44A2--,SMOX+,SMPD1-,SNAPIN-,SNHG12+,SNORA73A+,SNRNP40-,SOX12+,SPRED3-,SRM-,SRRM2+,STARD10+,SUMF1-,SUMF2-,SUOX-,SURF1-,SWI5-,TAB1-,TBC1D10A-,TEX264-,TFPT-,TIMELESS+,TKFC-,TM7SF2--,TMEM258-,TMPRSS5+,TNFSF13-,TPP1-,TRAPPC9-,TRIM47+,TRIM8+,TRPM4-,TRRAP+,TSHZ1+,TSR2-,TST-,TXN2--,UBE2I+,UQCR11-,URB1+,USP4-,VKORC1-,VRK3-,WNT4+,XYLT2-,ZBED6++,ZBTB7C+,ZDHHC11+,ZDHHC4--,ZMIZ1+,ZNF16-,ZNF219+,ZNF358+,ZNF445+,ZNF496-,ZNF500+,ZNF511+,ZNF589+,ZNF750+,ZNF768+,ZNF853+,ZNHIT1-,ZSCAN25- |
| **turquoise** | BP | GO:0006807 | nitrogen compound metabolic process | 1482 | 8098 | 6.78E-06 | 0.012 | AAAS-,AARS2-,ABHD14B-,ACAN+,ACOT8+,ACSS1-,ADAM15-,ADAM8-,ADAMTS13-,ADAMTS2++,ADPGK-,AGPAT1--,AKR7A2-,ALDH4A1-,ALYREF-,ANKRD9+,APH1A-,APLNR+,APOE-,ARHGEF10L+,ASPHD1-,ATAT1-,ATF5-,ATF6B-,ATG4B+,ATN1+,ATP5MC2-,AUTS2-,B3GALT4-,BCAP31-,BCKDHA-,BOLA1-,BRD1+,BRD2+,BRD4+,BRPF3+,CALR--,CASP9-,CBS+,CBY1-,CCDC88C-,CCS--,CHST6++,CIB1-,CIC+,CINP-,CLEC3B-,CNDP2-,COMT-,COQ9-,CRAT-,CREBBP+,CSNK2B-,CST3-,CSTB-,DCAF11-,DCAKD-,DDX49-,DELE1-,DESI1-,DGCR8+,DHX8-,DIDO1+,DLST-,DOK7+,DUS2-,EDEM2--,EEF2-,EEF2K+,EIF3D-,EIF5A-,ELOB-,FAH-,FANCE+,FBXL12-,FBXO17+,FBXO6-,FBXW8-,FGFR3-,FIS1-,FKBP2-,FKBP9-,FLAD1-,FLCN+,FN3KRP-,FSTL3+,G6PD-,GAMT-,GANAB-,GCDH-,GLI1+,GLMP-,GORASP1-,GPAT4+,GPR37L1-,GPX4-,GSTK1-,GSTP1-,GTF2H4-,GUK1-,H2BC4-,HDAC4++,HES4--,HES5-,HINT2--,HMBS-,HMGCL-,HNRNPL-,HR-,HRAS-,IKZF4+,IMP4-,INPP5K-,INTS3--,IRAK2++,IRF2BPL+,IRS2++,ITIH4-,IVD-,JADE2+,JARID2+,JDP2+,JUND+,KCTD5-,KDM4B+,KDM6B+,KLHL21+,LAMTOR1-,LARS2--,LSM7-,LTBP4+,MAPT-AS1+,MCM3-,MECP2+,MED20-,MFHAS1+,MGMT-,MICAL1-,MMP28--,MPV17L2-,MRM1-,MRPL20-,MRPL24--,MRPL53-,MRPS18B-,MSRB1--,MT3-,MTG2-,MYH3+,NCSTN-,NENF-,NFKBID+,NIT1-,NMRAL1-,NOL3-,NR6A1+,NTHL1-,NUCB1-,NUDT2--,NUP210-,OS9-,PARP3-,PARS2-,PC-,PDCD11-,PDE9A-,PER1+,PEX10-,PIM3+,PLK5+,PMM2-,PNPLA6+,POLR2G-,POLR2L-,PPIF-,PPP1CA-,PPP1R1B-,PPP4C-,PRELID1-,PRKCD--,PSENEN--,PSMB10-,PTPN7-,PTPRU+,PUF60-,RAD9A-,RASA4+,RBM10-,RECQL5-,RERE+,RFXANK-,RGMA+,RNF167-,RNF181-,RNF5-,RPN2-,RPS15-,RPS19-,RUVBL1-,S100A1--,SARDH-,SCARNA13+,SDSL--,SEC13--,SF3A1--,SF3B5-,SH2D3C-,SHISA5-,SKI+,SLC35C2-,SLC44A2--,SMOX+,SMPD1-,SNAPIN-,SNHG12+,SNORA73A+,SNRNP40-,SOX12+,SPRED3-,SRM-,SRRM2+,STARD10+,SUMF1-,SUMF2-,SUOX-,SURF1-,SWI5-,TAB1-,TBC1D10A-,TFPT-,TIMELESS+,TMEM258-,TMPRSS5+,TNFSF13-,TPP1-,TRAPPC9-,TRIM47+,TRIM8+,TRPM4-,TRRAP+,TSHZ1+,TSR2-,TST-,TXN2--,UBE2I+,UQCR11-,URB1+,USP4-,VKORC1-,VRK3-,WNT4+,XYLT2-,ZBED6++,ZBTB7C+,ZDHHC11+,ZDHHC4--,ZMIZ1+,ZNF16-,ZNF219+,ZNF358+,ZNF445+,ZNF496-,ZNF500+,ZNF511+,ZNF589+,ZNF750+,ZNF768+,ZNF853+,ZNHIT1-,ZSCAN25- |

| | | | | | | | | |
|---|---|---|---|---|---|---|---|---|
| **turquoise** | BP | GO:0071704 | organic substance metabolic process | 1583 | 8718 | 1.51E-05 | 0.017 | AAAS-,AARS2-,ABHD14B-,ACADS-,ACAN+,ACOT8+,ACSS1-,ADAM15-,ADAM8-,ADAMTS13-,ADAMTS2++,ADCYAP1R1+,ADIPOR1--,ADPGK-,AGPAT1--,AKR7A2-,ALDH4A1-,ALYREF-,ANKRD9+,APH1A-,APLNR+,APOE-,ARAP1+,ARHGEF10L+,ASPHD1-,ATAT1-,ATF5-,ATF6B-,ATG4B+,ATN1+,ATP5MC2-,AUTS2-,B3GALT4-,BCAP31-,BCKDHA-,BOLA1-,BRD1+,BRD2+,BRD4+,BRPF3+,BSG-,CACNA1H-,CALR--,CASP9-,CBS+,CBY1-,CCDC88C-,CCS--,CES2-,CHST6++,CIB1-,CIC+,CINP-,CLEC3B-,CNDP2-,COMT-,COQ9-,CRAT-,CREBBP+,CRYL1-,CSNK2B-,CST3-,CSTB-,CYB5R3-,DCAF11-,DCAKD-,DDX49-,DELE1-,DESI1-,DGCR8+,DHRS7B-,DHX8-,DIDO1+,DLST-,DOK7+,DUS2-,ECH1-,EDEM2--,EEF2-,EEF2K+,EIF3D-,EIF5A-,ELOB-,ETFB--,FAH-,FAM3A-,FANCE+,FBXL12-,FBXO17+,FBXO6-,FBXW8-,FGFR3-,FIS1-,FKBP2-,FKBP9-,FLAD1-,FLCN+,FMC1-,FN3KRP-,FSTL3+,G6PC3-,G6PD-,GAMT-,GANAB-,GCDH-,GLI1+,GLMP-,GORASP1-,GPAT4+,GPR37L1-,GPX4-,GSTK1-,GSTP1-,GTF2H4-,GUK1-,GYS1-,H2BC4-,HDAC4++,HES4--,HES5-,HINT2--,HMBS-,HMGCL-,HNRNPL-,HR-,HRAS-,IKZF4+,IMP4-,INPP5K-,INTS3--,IRAK2++,IRF2BPL+,IRS2++,ITIH4-,IVD-,JADE2+,JARID2+,JDP2+,JUND+,KCTD5-,KDM4B+,KDM6B+,KLHL21+,LAMTOR1-,LARS2--,LDHD-,LSM7-,LTBP4+,MAPT-AS1+,MCM3-,MECP2+,MED20-,MFHAS1+,MGMT-,MICAL1-,MMP28--,MPV17L2-,MRM1-,MRPL20-,MRPL24--,MRPL53-,MRPS18B-,MSRB1--,MT3-,MTG2-,MYH3+,NCSTN-,NENF-,NFKBID+,NMRAL1-,NOL3-,NR6A1+,NTHL1-,NUCB1-,NUDT2--,NUP210-,OS9-,PARP3-,PARS2-,PC-,PDCD11-,PDE9A-,PER1+,PEX10-,PIM3+,PLK5+,PLPP4+,PMM2-,PNPLA6+,POLR2G-,POLR2L-,PPIF-,PPP1CA-,PPP1R1B-,PPP4C-,PRELID1-,PRKCD--,PSENEN--,PSMB10-,PTPN7-,PTPRU+,PUF60-,RAD9A-,RASA4+,RBM10-,RECQL5-,RERE+,RETSAT-,RFXANK-,RGMA+,RNF167-,RNF181-,RNF5-,RPN2-,RPS15-,RPS19-,RUBCN+,RUVBL1-,S100A1--,SARDH-,SCARNA13+,SDSL--,SEC13--,SF3A1--,SF3B5-,SH2D3C-,SHISA5-,SKI+,SLC35C2-,SLC44A2--,SMOX+,SMPD1-,SNAPIN-,SNHG12+,SNORA73A+,SNRNP40-,SOX12+,SPRED3-,SRM-,SRRM2+,STARD10+,SUMF1-,SUMF2-,SUOX-,SURF1-,SWI5-,TAB1-,TBC1D10A-,TFPT-,TIMELESS+,TKFC-,TM7SF2--,TMEM258-,TMPRSS5+,TNFSF13-,TPP1-,TRAPPC9-,TRIM47+,TRIM8+,TRPM4-,TRRAP+,TSHZ1+,TSR2-,TST-,TXN2--,UBE2I+,UQCR11-,URB1+,USP4-,VKORC1-,VRK3-,WNT4+,XYLT2-,ZBED6++,ZBTB7C+,ZDHHC11+,ZDHHC4--,ZMIZ1+,ZNF16-,ZNF219+,ZNF358+,ZNF445+,ZNF496-,ZNF500+,ZNF511+,ZNF589+,ZNF750+,ZNF768+,ZNF853+,ZNHIT1-,ZSCAN25- |
| **turquoise** | BP | GO:0006325 | chromatin organization | 153 | 659 | 1.57E-05 | 0.017 | AUTS2-,BRD1+,BRD2+,BRD4+,BRPF3+,CREBBP+,FLCN+,GPX4-,H2BC18-,H2BC4-,HDAC4++,HR-,JADE2+,JARID2+,JDP2+,KDM4B+,KDM6B+,MECP2+,MT3-,PER1+,PHF13+,PRKCD--,RERE+,RUVBL1-,SKI+,TRRAP+,ZMIZ1+,ZNHIT1- |
| **turquoise** | BP | GO:0043543 | protein acylation | 63 | 222 | 1.65E-05 | 0.017 | ATAT1-,AUTS2-,BRD1+,BRPF3+,CREBBP+,DLST-,FLCN+,HINT2--,JADE2+,MECP2+,PER1+,RUVBL1-,TRRAP+,ZDHHC11+,ZDHHC4-- |
| **turquoise** | BP | GO:0043170 | macromolecule metabolic process | 1375 | 7509 | 2.30E-05 | 0.021 | AAAS-,AARS2-,ABHD14B-,ACAN+,ACOT8+,ADAM15-,ADAM8-,ADAMTS13-,ADAMTS2++,ALYREF-,ANKRD9+,APH1A-,APLNR+,APOE-,ARAP1+,ARHGEF10L+,ASPHD1-,ATAT1-,ATF5-,ATF6B-,ATG4B+,ATN1+,AUTS2-,B3GALT4-,BCAP31-,BOLA1-,BRD1+,BRD2+,BRD4+,BRPF3+,CALR--,CASP9-,CBS+,CBY1-,CCDC88C-,CCS--,CHST6++,CIB1-,CIC+,CINP-,CLEC3B-,CNDP2-,CREBBP+,CSNK2B-,CST3-,CSTB-,DCAF11-,DDX49-,DELE1-,DESI1-,DGCR8+,DHX8-,DIDO1+,DLST-,DOK7+,DUS2-,EDEM2--,EEF2-,EEF2K+,EIF3D-,EIF5A-,ELOB-,FANCE+,FBXL12-,FBXO17+,FBXO6-,FBXW8-,FGFR3-,FIS1-,FKBP2-,FKBP9-,FLCN+,FN3KRP-,FSTL3+,G6PD-,GANAB-,GLI1+,GLMP-,GORASP1-,GPR37L1-,GSTP1-,GTF2H4-,GYS1-,H2BC4-,HDAC4++,HES4--,HES5-,HINT2--,HMBS-,HNRNPL-,HR-,HRAS-,IKZF4+,IMP4-,INPP5K-,INTS3--,IRAK2++,IRF2BPL+,IRS2++,ITIH4-,JADE2+,JARID2+,JDP2+,JUND+,KCTD5-,KDM4B+,KDM6B+,KLHL21+,LAMTOR1-,LARS2--,LSM7-,LTBP4+,MAPT-AS1+,MCM3-,MECP2+,MED20-,MFHAS1+,MGMT-,MICAL1-,MMP28--,MPV17L2-,MRM1-,MRPL20-,MRPL24--,MRPL53-,MRPS18B-,MSRB1--,MT3-,MTG2-,MYH3+,NCSTN-,NENF-,NFKBID+,NMRAL1-,NOL3-,NR6A1+,NTHL1-,NUCB1-,NUP210-,OS9-,PARP3-,PARS2-,PC-,PDCD11-,PER1+,PEX10-,PIM3+,PLK5+,PMM2-,POLR2G-,POLR2L-,PPIF-,PPP1CA-,PPP1R1B-,PPP4C-,PRELID1- |

| | | | | | | | | |
|---|---|---|---|---|---|---|---|---|
| | | | | | | | | ,PRKCD--,PSENEN--,PSMB10-,PTPN7-,PTPRU+,PUF60-,RAD9A-,RASA4+,RBM10-,RECQL5-,RERE+,RFXANK-,RGMA+,RNF167-,RNF181-,RNF5-,RPN2-,RPS15-,RPS19-,RUVBL1-,S100A1--,SCARNA13+,SEC13--,SF3A1--,SF3B5-,SH2D3C-,SHISA5-,SKI+,SLC35C2-,SMPD1-,SNAPIN-,SNHG12+,SNORA73A+,SNRNP40-,SOX12+,SPRED3-,SRRM2+,SUMF1-,SUMF2-,SWI5-,TAB1-,TBC1D10A-,TFPT-,TIMELESS+,TMEM258-,TMPRSS5+,TNFSF13-,TPP1-,TRAPPC9-,TRIM47+,TRIM8+,TRPM4-,TRRAP+,TSHZ1+,TSR2-,UBE2I+,URB1+,USP4-,VKORC1-,VRK3-,WNT4+,XYLT2-,ZBED6++,ZBTB7C+,ZDHHC11+,ZDHHC4--,ZMIZ1+,ZNF16-,ZNF219+,ZNF358+,ZNF445+,ZNF496-,ZNF500+,ZNF511+,ZNF589+,ZNF750+,ZNF768+,ZNF853+,ZNHIT1-,ZSCAN25- |
| **turquoise** | BP | GO:0016569 | covalent chromatin modification | 103 | 419 | 3.71E-05 | 0.030 | AUTS2-,BRD1+,BRD4+,BRPF3+,CREBBP+,FLCN+,HDAC4++,HR-,JADE2+,JARID2+,JDP2+,KDM4B+,KDM6B+,MECP2+,MT3-,PER1+,PRKCD--,RUVBL1-,SKI+,TRRAP+,ZNHIT1- |
| **turquoise** | MF | GO:0003682 | chromatin binding | 109 | 450 | 4.08E-05 | 0.030 | ATF5-,AUTS2-,BRD2+,BRD4+,CIC+,CREBBP+,CSNK2B-,GLI1+,HDAC4++,HES5-,HR-,JARID2+,JDP2+,KDM6B+,PER1+,PHF13+,RERE+,TSHZ1+,ZNF750+,ZNHIT1- |
